# Clinical Validation of Saliency Maps for Understanding Deep Neural Networks in Ophthalmology

**DOI:** 10.1101/2021.05.05.21256683

**Authors:** Murat Seçkin Ayhan, Louis Benedikt Kümmerle, Laura Kühlewein, Werner Inhoffen, Gulnar Aliyeva, Focke Ziemssen, Philipp Berens

## Abstract

Deep neural networks (DNNs) have achieved physician-level accuracy on many imaging-based medical diagnostic tasks, for example classification of retinal images in ophthalmology. However, their decision mechanisms are often considered impenetrable leading to a lack of trust by clinicians and patients. To alleviate this issue, a range of explanation methods have been proposed to expose the inner workings of DNNs leading to their decisions. For imaging-based tasks, this is often achieved via saliency maps. The quality of these maps are typically evaluated via perturbation analysis without experts involved. To facilitate the adoption and success of such automated systems, however, it is crucial to validate saliency maps against clinicians. In this study, we used three different network architectures and developed ensembles of DNNs to detect diabetic retinopathy and neovascular age-related macular degeneration from retinal fundus images and optical coherence tomography scans, respectively. We used a variety of explanation methods and obtained a comprehensive set of saliency maps for explaining the ensemble-based diagnostic decisions. Then, we systematically validated saliency maps against clinicians through two main analyses — a direct comparison of saliency maps with the expert annotations of disease-specific pathologies and perturbation analyses using also expert annotations as saliency maps. We found the choice of DNN architecture and explanation method to significantly influence the quality of saliency maps. Guided Backprop showed consistently good performance across disease scenarios and DNN architectures, suggesting that it provides a suitable starting point for explaining the decisions of DNNs on retinal images.

## Introduction

Deep neural networks (DNNs) have become increasingly popular in medical image analysis [50, 26, 89, 73, 27]. Trained on various diagnostic tasks in imaging-based specialties of medicine, they have been shown to achieve physician-level accuracy [35, 25, 20, 37, 7, 94]. However, DNNs are often referred to as *black boxes* since their decision mechanisms are not transparent enough for clinicians to interpret and trust them [16, 52, 73]. From a practical and ethical point of view, this is one of the major roadblocks in translating cutting-edge machine learning research into meaningful clinical tools [28, 33, 34]. To tackle this challenge, a number of explanation methods have been proposed to expose the inner workings of a DNN underlying its decisions. In the case of image analysis, this is frequently done via *saliency maps*, where input pixels are associated with saliency scores according to their contribution to network outputs [4, 60]. The efficacy of a saliency map is typically evaluated via perturbation or sensitivity analysis [11, 76, 4, 44, 60, 62], without involving a human in the process. For medical imaging, we thus lack an understanding of how good different explanation methods are in providing saliency maps with clinical relevance.

To fill this gap, we systematically evaluated saliency maps for the decisions of DNNs trained to detect two prevalent eye diseases, diabetic retinopathy (DR) and neovascular age-related macular degeneration (nAMD), with respect to the expert opinions of clinical ophthalmologists. First, we compared saliency maps with disease-specific annotations of pathologies. Second, we performed perturbation analyses and compared the outcome to that obtained when using expert annotations as saliency maps. This allowed us to use perturbation analysis also as a tool to validate DNN explanations against clinicians.

We also introduced two technical novelties. First, we developed a post-processing method for saliency maps to improve the visualization of salient regions and standardize saliency maps for benchmarks. In addition, we computed saliency for *Deep Ensembles* [47, 29] to obtain saliency maps which were more informed than those obtained from individual networks in isolation.

## Methods

DNNs are often trained for diagnostic classification of medical images. To introduce notation, we first review the basics of DNN-based image classification. Then, we describe our datasets and disease detection tasks as well as our methodology including model development and evaluation. Also, we discuss attribution methods for generating saliency maps and introduce our post-processing method within this classification framework.

In medical image analysis, a DNN achieves a diagnostic classification by learning a function that map inputs to outputs: *y* = *f*_*θ*_(**x**), where *y* is a class label (e.g. disease severity or presence/absence of disease) assigned by experts to an input image **x** and *θ* represents the DNN’s weights, which are tuned w.r.t. an objective on a finite dataset 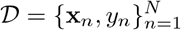. The objective is usually to minimize the cross-entropy between labels and predictions, which can be expressed in the following form: 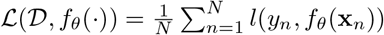, Where 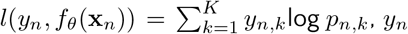 is a hard label in multinomial (1-hot) representation, *p*_*n*_ is a list of predicted class probabilities and *k* is an index into *K* classes. A DNN estimates the class probabilities typically via a *softmax* function in its final layer: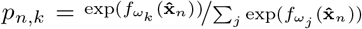, where *ω*_*k*_ ⊂ *θ* represents the weights and bias for the *k*-th class in the softmax layer, 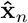 is the feature representation by the network’s penultimate layer, and outputs are multinomial distributions: ∑ _*k*_ *p*_*n,k*_ = 1.

## Diseases and Datasets

DR and nAMD are two prevalent and progressive eye diseases [18, 3, 95], both of which can be automatically graded using state-of-the-art DNNs [35, 20, 77, 46, 94].

In the case of DR, we used multiple publicly available collections of fundus images (Fig. 1a): Kaggle DR [41], Asia Pacific Tele-Ophthalmology Society (APTOS) DR [42], Messidor 2 [1, 21, 45], and Indian Diabetic Retinopathy Image Dataset (IDRiD) [70]. These images are graded by medical experts according to the International Clinical Diabetic Retinopathy Severity Scale (Table 1). In addition to the image-level DR grades, 81 of the IDRiD images are annotated at the pixel level with regards to pathologies associated with DR, i.e., microaneurysms, soft exudates, hard exudates and hemorrhages as well as the optic disc [70] (Fig. 1b).

**Table 1:**
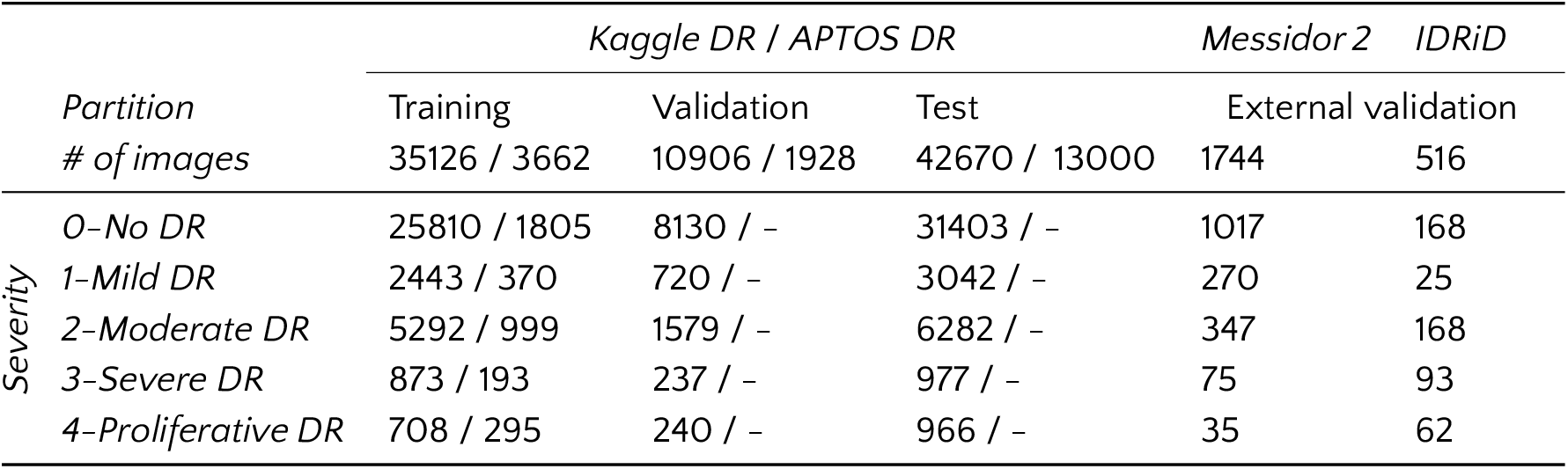
Fundus image collections. Kaggle DR and APTOS DR partitions are given according to the source. Unpublished labels are indicated with ’-’. Messidor 2 and IDRiD data are used for external validation only.

**Figure 1:**
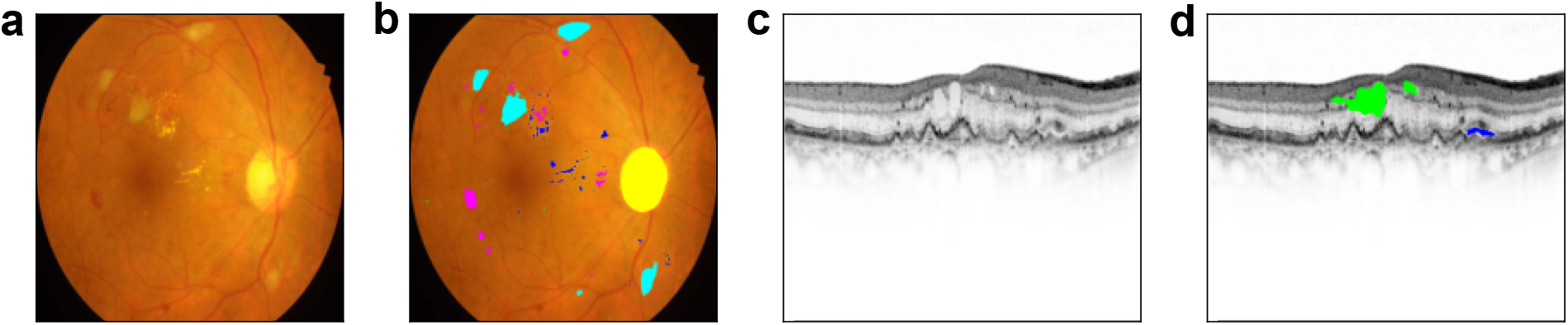
Exemplary retinal images along with their pixel-level annotations for lesions. Best viewed in color and when zoomed in. **(a)**: A fundus image from the IDRiD collection. **(b)**: The fundus image with the annotations for microaneurysms (green), hemorrhages (magenta), hard exudates (blue), soft exudates (cyan) and the optic disc (yellow). **(c)**: A B-scan from our OCT collection. **(d)**: The same B-scan annotated for retinal fluid. Intraretinal fluid is marked by green, whereas blue indicates subretinal fluid.

In the case ofnAMD, we used 70 3D optical coherence tomography (OCT) volume scans from patients at the University Eye Hospital Tübingen collected with Heidelberg Spectralis OCT (Heidelberg Engineering, Heidelberg, Germany). Depending on the settings used during clinical examinations, each volume consisted of 19,25,37,49 or 73 2D slices, namely B-scans (Fig. 1c). In total, there were 3762 B-scans (1751 left eye, 2011 right eye) and each was graded by a retina specialist according to the presence or absence of *active* nAMD (Table 2), which is characterized by intraretinal or subretinal fluid (Fig. 1d). Furthermore, we selected 73 B-scans from the validation (19) or test (54) sets to be annotated by a board-certified ophthalmologist at the pixel level w.r.t. nAMD activity. We excluded two of the annotated B-scans from our analyses due to the mismatch between their image-level grading and pixel-level annotations carried out by our clinicians (WI and LK, respectively). The use of this data set was permitted by the Institutional Ethics Committee of the University of Tübingen and was performed in line with all relevant laws and regulations.

**Table 2:** OCT collection and B-scans.

| Partition | Training | Validation | Test |
| --- | --- | --- | --- |
| # of patients | 53 | 7 | 10 |
| # of B-scans | 2751 | 407 | 604 |
| *0-Inactive* | 1903 | 306 | 335 |
| *1-Active* | 848 | 101 | 269 |

## Diagnostic Tasks, Network Architectures and Model Development

In the DR case, the task for the network was to detect DR from fundus images. Considering Mild DR as the disease onset (Table 1), we grouped the fundus images into *healthy* and *diseased*, according to the DR stages: {0} vs. {1,2,3,4}, respectively. For the nAMD case, the task was to recognize the nAMD activity from individual B-scans of the retina (Table 2). For both tasks, we mainly used two well-established DNN architectures, ResNet50 [38] and InceptionV3 [86]. We also adopted EfficientNets [87] to obtain insights into newer architectures. We obtained the ResNet50 and InceptionV3 implementations from Keras [19], and EfficientNet’s from a public repository [92] also based on Keras. All networks were pretrained on ImageNet [75]. We modified and fine-tuned them to our tasks. Also, we used a 2-way softmax encoding for the classification outcome for the sake of compatibility with the saliency methods (see Explanations in the Visual Domain).

### DR detection networks

The pretrained networks originally included 1000-way softmax for classification on ImageNet. We modified the classification layer in each network by adding a new dense layer with 512 units followed by Batch Normalization [40] and ReLU [63]. Then, we used a simple 2-way softmax layer. We applied *L*_2_ and *L*_1_ regularizers to convolutional and penultimate layers, respectively. Also, we modified the objective functions to handle the class imbalance in the datasets (Table 1): 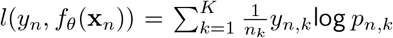, where *n*_*k*_ is the number of images from class *k* in a minibatch. Using Stochastic Gradient Descent (SGD) with Nesterov’s Accelerated Gradients (NAG) [64, 85] and a momentum coefficient of 0.9, we trained the networks for 150 epochs on random partitions of all labeled images from *Kaggle DR* and *APTOS DR* combined (92,364 images, Table 1). More specifically, we performed 5-fold cross-validation within these images and used 80% of them for training. For each cross-validation run, we followed a stepwise learning rate schedule with rates 0.005, 0.001, 0.0005, 0.0001 after epochs 0, 25, 50, 85 respectively, on top of a decay rate of 0.00001. Also, during the first 10 epochs, only the dense layers were updated and convolutional layers were frozen. For the remaining epochs, all layers were fine-tuned to the task. The model performance was validated after each epoch on the remaining 20% of the images and the best configuration was saved for inference. In this scheme, each DNN instance was evaluated on a disjoint *internal* validation set. In order to get a better picture of our DNNs’ generalization performance, we finally evaluated them on an *external* validation set that comprised of both Messidor 2 and IDRiD images (Table 1).

### nAMD activity detection networks

We modified the pretrained networks by concatenating max pooling to average pooling, adding two dense layers with 1024 and 512 units, which were also followed by Batch Normalization [40] and ReLU activation [63], and using a 2-way softmax classifier. The use of both max and average pooling led to performance improvements in our previous work [49, 9, 10]. Also, all weight layers except the penultimate one were equipped with *L*_2_ regularization. We used *L*_1_ regularization to promote sparsity in the penultimate layer. Ultimately, all networks achieved classification based on 512 features obtained from their penultimate layers. In this case, we countered the class imbalance (Table 2) with random oversampling. Using SGD with NAG [64, 85], a momentum coefficient of 0.9, initial learning rate of 0.001, a decay rate of 0.0001 and a regularization constant of 0.00001, we trained networks for 100 epochs. During the first 10 epochs, the convolutional stacks were frozen and only the dense layers were trained. For the remaining epochs, all layers were fine-tuned to the task. The best models based on validation accuracy were saved after each epoch and used for inference on the test set.

### Data augmentation and image preprocessing

Fundus images exhibited various sizes. Their dimensions typically varied in [1900, 4800] pixels, with width being larger than height. Thus, fundus images were first cropped to center such that the fundus circle touched the image borders. Namely, the longer axis of image height or width was cropped on both sides equally to the same length as the shorter axis. Then, images were resized to 512 *×* 512. During training, data augmentation was applied to the images. The augmentation pipeline included random operations: vertical and horizontal flips, rotation within *±*180 degrees (pixels that have no image information due to rotation were set to black pixels), horizontal and vertical translations within *±*20 pixels, brightness adjustments within *±*30% and zoom within [−20%, 0%]. After the first preprocessing and data augmentation, the specific preprocessing functions of ResNet50 or InceptionV3 from the Keras API [19] were applied. For EfficientNets, the preprocessing function was from the aforementioned repository [92].

B-scans contained 440 *×* 512 pixels (Fig. 1c). We performed data augmentation before feeding images to networks during training. The augmentation pipeline included random rotation within *±*45 degrees, horizontal and vertical translations within *±*30 pixels, brightness adjustments within *±*10%, zoom within *±*10%, and horizontal and vertical flips. Once images went through the pipeline, they were locally color-normalized for contrast enhancement with background subtraction via a median filter of size 31. Then, appropriate preprocessing functions from the Keras API [19] or the EfficientNet repository [92] were applied.

### Overconfidence and calibration of predictive probabilities via Deep Ensembles

DNNs are overconfident about their predictions [36, 90, 55]. Their predictive probabilities do not reflect the true probability of the predictions being correct, namely accuracy. Such probabilistic outputs are said to be *miscalibrated* and they do not lead to well-calibrated, reliable uncertainty estimates regarding DNNs’ decisions [30, 36, 43, 47, 54, 22, 90]. To obtain well-calibrated predictions and improve the performance of our networks, we used Deep Ensembles [47]. A Deep Ensemble simply consists of multiple DNNs, each of which is randomly initialized, follows a different optimization trajectory and explores a different mode in function space [47, 29]. Thus, the ensemble, even a small one with 3-5 DNNs, samples diverse and accurate predictors from a function space, exploits their diversity in decision-making and ultimately improves upon the single network performance both in accuracy and calibration [47, 29, 68], also in a DR detection scenario [10]. Using the network architectures, hyperparameters and training procedures described above, we constructed ensembles of 5 DNNs for our diagnostic tasks (Table 3, Fig. 2). In the DR case, we used the DNNs trained during cross-validation. For the nAMD task, we trained 5 DNNs per architecture. All DNNs were diversified by the randomness in the initialization of dense layers, shuffling of training examples as well as data augmentation.

**Table 3:**
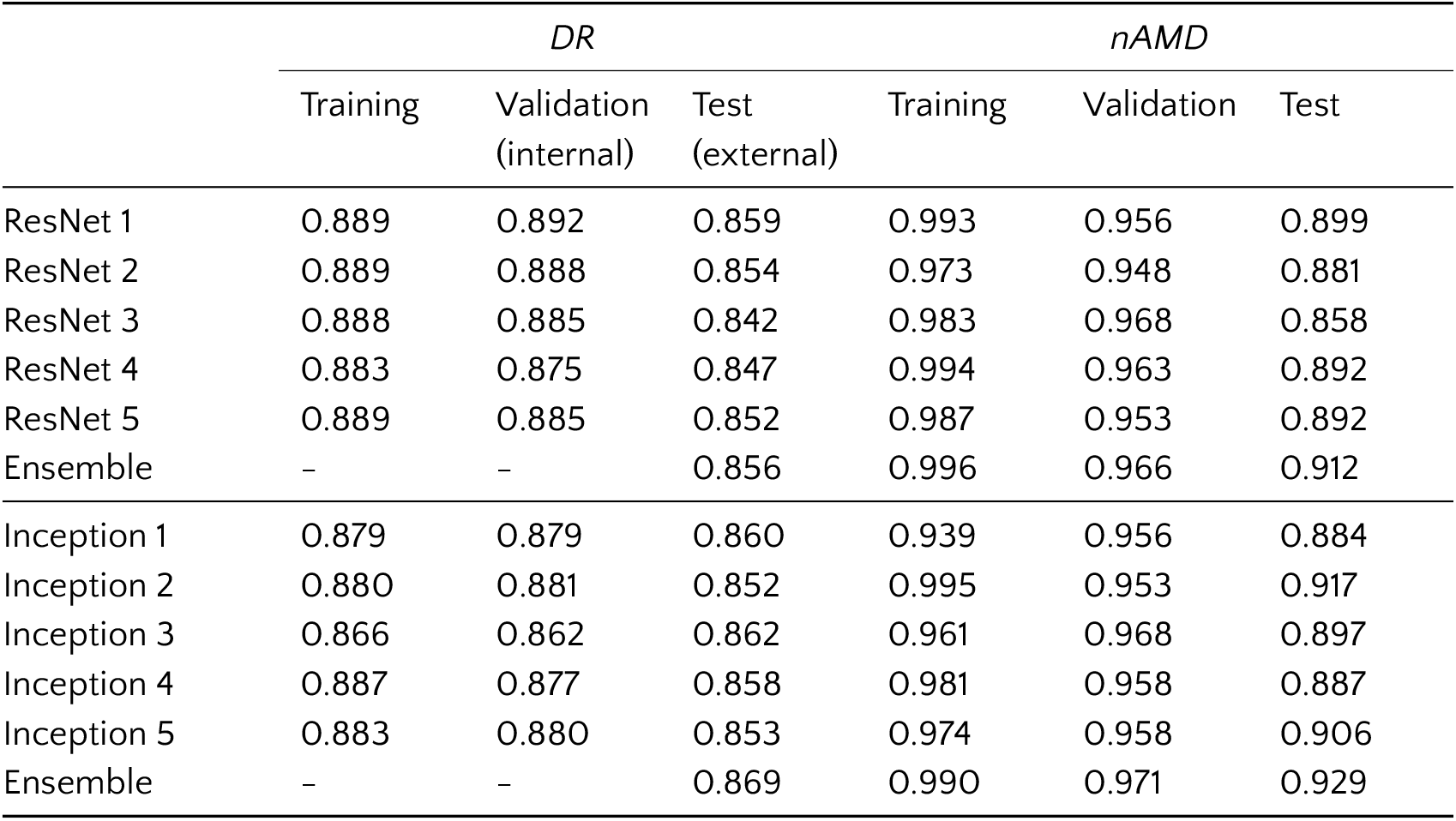
Disease detection accuracy for individual networks and their ensembles. For EfficientNets, see Appendix A.5.

|  | *DR* |  |  | *nAMD* |  |  |
| --- | --- | --- | --- | --- | --- | --- |
|  | Training | Validation<br>(internal) | Test<br>(external) | Training | Validation | Test |
| ResNet 1 | 0.889 | 0.892 | 0.859 | 0.993 | 0.956 | 0.899 |
| ResNet 2 | 0.889 | 0.888 | 0.854 | 0.973 | 0.948 | 0.881 |
| ResNet 3 | 0.888 | 0.885 | 0.842 | 0.983 | 0.968 | 0.858 |
| ResNet 4 | 0.883 | 0.875 | 0.847 | 0.994 | 0.963 | 0.892 |
| ResNet 5 | 0.889 | 0.885 | 0.852 | 0.987 | 0.953 | 0.892 |
| Ensemble | – | – | 0.856 | 0.996 | 0.966 | 0.912 |
| Inception 1 | 0.879 | 0.879 | 0.860 | 0.939 | 0.956 | 0.884 |
| Inception 2 | 0.880 | 0.881 | 0.852 | 0.995 | 0.953 | 0.917 |
| Inception 3 | 0.866 | 0.862 | 0.862 | 0.961 | 0.968 | 0.897 |
| Inception 4 | 0.887 | 0.877 | 0.858 | 0.981 | 0.958 | 0.887 |
| Inception 5 | 0.883 | 0.880 | 0.853 | 0.974 | 0.958 | 0.906 |
| Ensemble | – | – | 0.869 | 0.990 | 0.971 | 0.929 |

**Figure 2:**
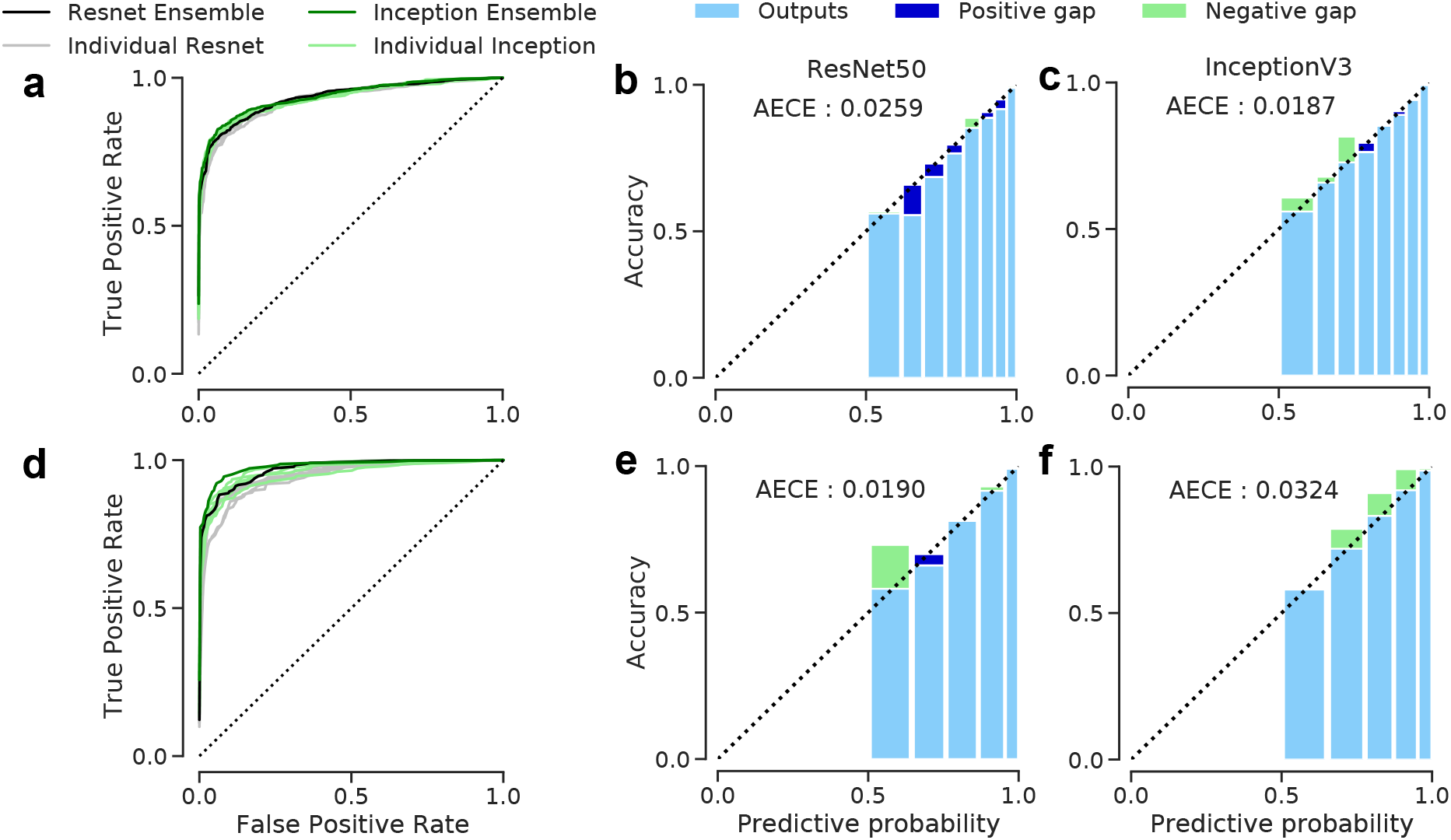
Receiver Operating Characteristics (ROCs) and calibration of our ensembles. The degree of mis-calibration was estimated via reliability diagrams [22, 36, 65] and the Adaptive Expected Calibration Error (AECE) [22] based on adaptive histograms. A positive gap (dark blue) between predictive probability and accuracy indicates overconfidence, whereas a negative gap (light green) points at the lack of confidence. **(a-c)**: DR detection. For the sake of clarity, only the performances on external validation set are shown. **(d-f)**: nAMD activity detection. Only the test set performances are shown.

### Explanations in the Visual Domain

Saliency maps are frequently used to obtain explanations for a DNN’s decisions. We focused on saliency methods with implicit access to model structure and its internal state. These methods generate saliency maps via forward and backward passes [71, 4, 60, 62, 73]. They typically use backpropagation-based algorithms or relevance propagation rules. As a result, a DNN’s decision is unravelled by attributing its predictive values all the way back to the input domain [4, 60, 62, 73]. In this sense, an attribution is a mapping *h* from an RGB image **x** to its raw saliency map through a trained network with *K* outputs.

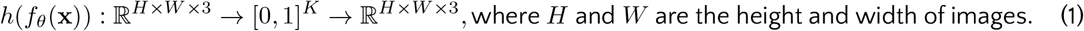

In order to compute saliency maps conveniently, we used the open-source library *iNNvestigate* [2]. We only considered common gradient or relevance-based methods, which included a variety of methods commonly used in ophthalmology and neuroimaging [74, 6, 5, 14, 77, 59, 93].

### Gradient-based methods

A saliency map *R* for an image can be obtained by simply using backpropagation to compute the gradient of the predictive function w.r.t. inputs indexed by *i*, given a class of interest 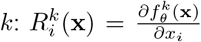 [80, 60]. However, gradients are sensitive to pixel-based variation and yield scattered saliency maps [60, 66]. To reduces this sensitivity, Simple Taylor decomposition [11], which is also called input *×* gradient, emphasizes an input only if it is present and the network responds to it [60]: 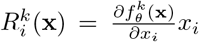. Also, Deep Taylor decomposition (DTD) [61] computes the relevance scores in a layer-wise fashion: 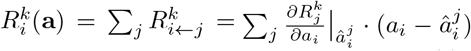, where *j* is an index into connections, **a** represents activations and â is a root point used in decomposition. Here, the *i*-th neuron in a given layer receives relevance scores from its connections to the next layer w.r.t. derivative evaluations at 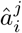. DTD also ensures the positivity of relevance scores at each layer through local decompositions and constraints [61].

SmoothGrad [82] reduces the pixel-sensitivity of gradients by sampling inputs with additive noise and averaging over multiple maps. Its goal is to generate more informed and focused maps. Similarly, Integrated Gradients [84] assumes a baseline (blank) image 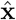 and follows a path between the baseline and input 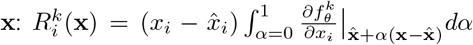 The gradients are integrated along the path. In practice, this means an approximation with a number of steps (e.g., 20-300 [84]) between **x** and 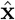. These sampling-based methods induce high computational costs, when large samples are needed for accurate explanations. Despite the cost, we used 256 samples (or steps) for the sake of accuracy, unless stated otherwise.

Apart from using the model structure as is, DeConvNet [97, 96] reverses the network components, e.g., pooling layers, filters and activations, and maps high-level features to inputs. In addition to deconvolution, Guided Backprop [83] resorts to a combination of both forward and backward ReLUs during backpropagation for sharper visualization [66, 74, 14]. However, it is restricted to ReLU networks, such as ResNet50 and InceptionV3.

### Layer-wise Relevance Propagation (LRP)

LRP [11] also relies on backward propagation but its *conservation principle* sets it apart from gradient-based methods. Within the LRP framework, each neuron distributes to its predecessors exactly the sum of relevance scores it receives from its successors [61, 60, 62]. As a result, an *unnormalized* network output(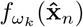, namely *logit*) reaches the input layer and disseminates into saliency scores. In this regard, LRP explains the actual predictive outputs, instead of their local variation. It supports both positive and negative relevance, corresponding to the excitation or inhibition characteristics of neurons, respectively [61, 60, 62].

A simple propagation rule is the *z*-rule (LRP-Z or LRP-0): 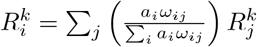, where *ω ⊂ θ* between two layers. LRP-*ε* introduces an additional hyperparameter *ε* to suppress the impact of weak or noisy contributions from successors [62]: 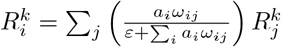 We defaulted to *ε* = 0.05. A general rule is the *αβ*-rule [61, 60, 62]: 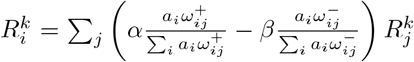, where *α* − *β* =1, *β* > 0, ^+^ and ^−^ denote the excitatory and inhibitory parts. The hyperparameters *α* and *β* set the balance between the positive and negative relevance and modulate the behaviour of saliency maps. Thanks to the conservation principle, more sophisticated rules can also be composed of simple ones. For instance, LRP-*αβ* can modulate the flow of relevance through the convolutional layers, while LRP-*ε* emphasizes the most salient scores through the dense layers [62]. We considered two such rules designated as LRP-PresetA and LRP-PresetB with *α* = 1, *β* = 0 and *α* = 2, *β* = 1, respectively [2]. These can also be coupled with a *flat* rule that assumes uniform weights, i.e., *ω* = 1, in the very first layer during the propagation of relevance. As a result, the sensitivity to the first layer convolutional filters is reduced and the effect of higher layers is emphasized.

### Post-processing and ensembling of saliency maps

Saliency maps essentially highlight regions in images based on which DNNs make their decisions. Thus, we summarized the raw saliency maps (see Eq. 1) into 2D, by summing up the saliency scores along channels. Then, we dispatched the positive and negative scores back into the *red* and *blue* channels, respectively, for visualization of excitatory or inhibitory features (Fig. 3). As the saliency scores exhibited stark differences due to the underlying assumptions and objectives of attribution methods, we mapped the absolute values of total scores into [0, 1] within channels. However, a naïve mapping via min-max normalization led to extremely sparse maps, even with ensembling (Fig. 3, last column) and various attribution methods (Fig. 5, top rows in (a) and (b)). We proposed a *non-linear* transformation to improve the visualization of salient regions. Our procedure is a *drop-in* replacement for the min-max normalization.

**Figure 3:**
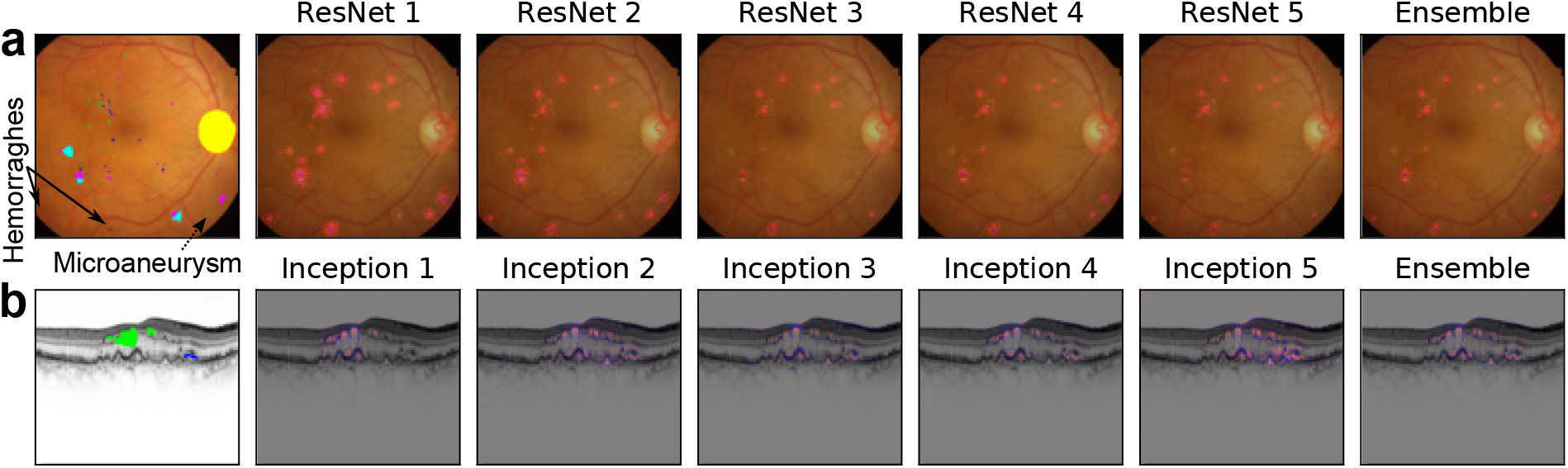
Post-processing and ensembling of saliency maps. All models correctly predicted the presence of DR or active nAMD, given the inputs. Coloring of annotations is the same as in Fig. 1. Raw saliency scores were obtained via Guided Backprop [83], aggregated along channels, positive and negative scores were separated into the red and blue channels, respectively, and their absolute values were min-max normalized into [0, 1] within channels. Then, ensemble-based maps were obtained by simple averaging. Best viewed in color and when zoomed in. **(a)** Exemplary saliency maps from the ResNet50 instances and their ensemble for DR detection. **(b)** Same as (a) but with InceptionV3 instances and for the nAMD activity detection.

Given a 2D map 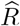 for excitatory or inhibitory features, we rescaled its values w.r.t. the maximum possible sum of scores the map could have had after processing, i.e., 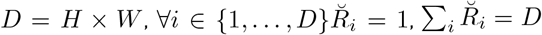 :

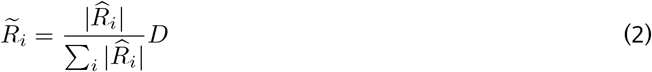

Then, we achieved a non-linear transformation by thresholding and another rescaling (Fig. 4):

**Figure 4:**
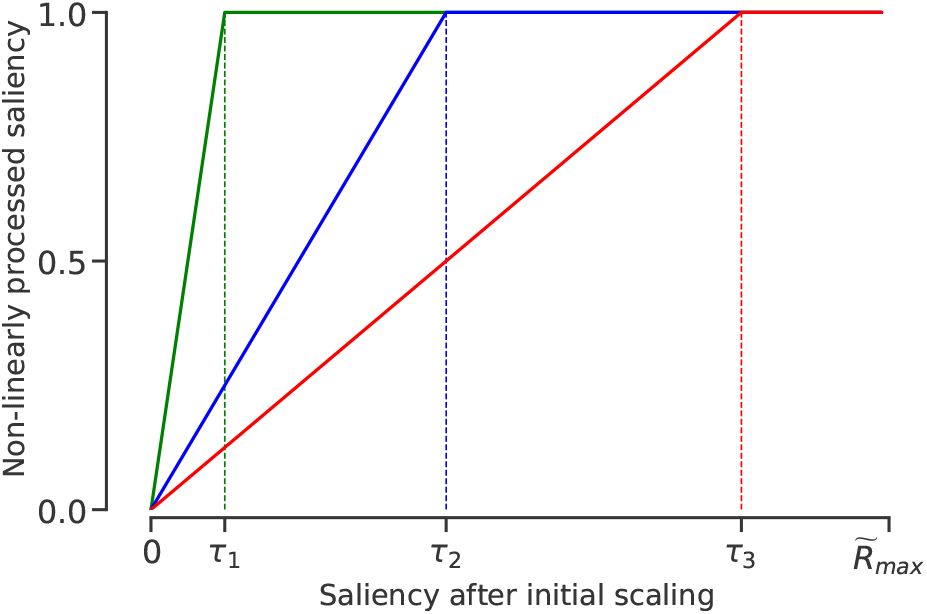
Illustration of our saliency map processing via thresholding and scaling. Given a threshold *τ*, saliency scores above the threshold are clipped to 1 and others are linearly scaled between 0 and 1.

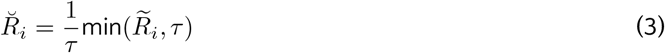

We determined the threshold *τ* by solving the following problem:

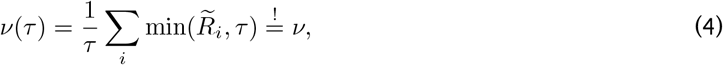

where *ν* was our target for total relevance and *ν*(*τ*) was a monotonically decreasing and implicit function of 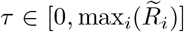 (see Appendix A.1) with upper and lower bounds: lim _τ → 0_ *ν*(*τ*) *D* and 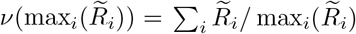, respectively. We performed a binary search to find a suitable *τ*. We also introduced a hyperparameter *f*_*ν*_ so that *ν* was easily adjusted: *ν* = *f*_*ν*_ *D*, where *f*_*ν*_ *∈* [0, 1] was the fraction of *D*. Intuitively, *f*_*ν*_ allowed us to grow salient regions for better visualization (Fig. 5). However, the size of salient regions also depended on disease status and total class evidence carried over to logits. To update our initial choice in the light of evidence, we introduced a scaling parameter:

**Figure 5:**
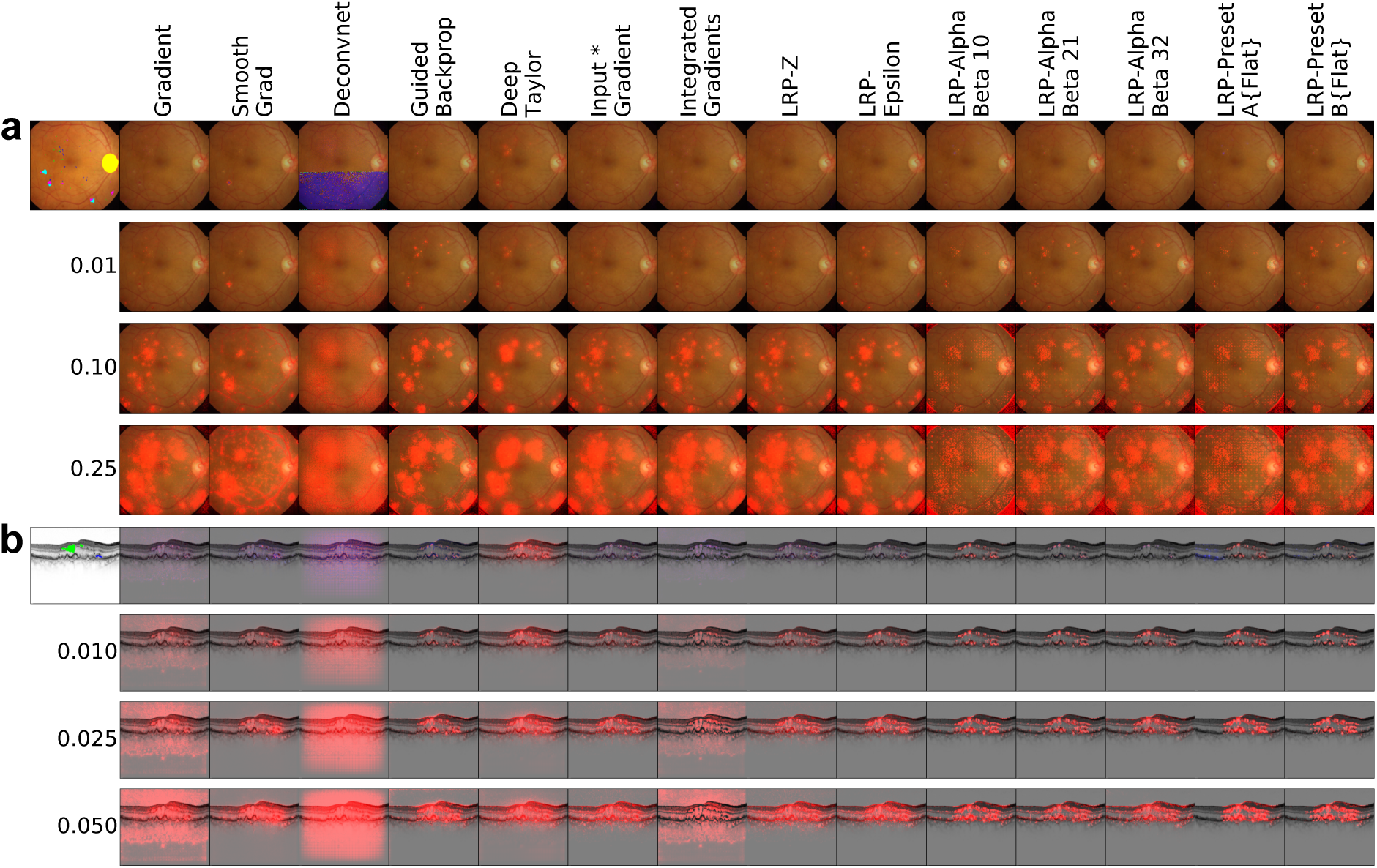
The impact of our post-processing on saliency maps. Top rows in **(a)** and **(b)** show annotated images (leftmost column) and saliency maps obtained via 14 attribution methods from the InceptionV3 ensembles using the min-max normalization as in Fig. 3. Remaining rows show the results of our post-processing w.r.t. various settings of *f*_*ν*_. **(a)** Exemplary saliency maps obtained from the DR detection ensemble for its prediction on the given fundus image, also post-processed w.r.t. 3 values of *f*_*ν*_ : 0.01, 0.1 and 0.25. Also note that we could not couple the LRP-PresetA and LRP-PresetB rules with the flat rule due to numerical difficulties. **(b)** Exemplary saliency maps obtained from the nAMD activity detection ensemble for its prediction on the given B-scan, also post-processed w.r.t. 3 values of *f*_*ν*_ : 0.01, 0.025 and 0.05.

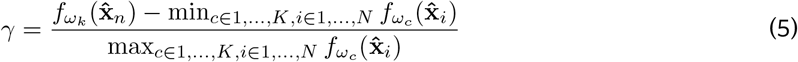

where the evidence 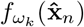 for class *k*, given an input image **x**_*n*_, was rescaled into [0, 1] w.r.t minimum and maximum evidence over all images and across classes. Then, *ν* = *γf*_*ν*_ *D*, which allowed for fine-tuning the ratios of salient regions with disease patterns and regions without over the image size. If the search interval was somehow violated after these adjustments, then we set 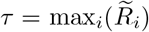 as a precaution. Also, in order to avoid *τ* = 0, we heuristically set the minimum possible *τ* to 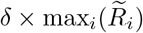, where *δ* = 0.00001.

In principle, our method could be used on saliency maps with both excitatory and inhibitory features. However, we focused on the excitatory ones since our evaluation was concerned with the efficacy of saliency maps for explaining the DNN decisions in presence of lesions and their annotations by clinicians. In addition, we leveraged the local sensitivity of gradient-based methods in order to enhance their visualizations of salient regions. Namely, we took the absolute values of raw saliency scores beforehand, which was a handy trick used for Guided Backprop in recent applications [74, 14]. Given the similarities between gradient-based saliency maps and those from LRP-Z and LRP-Epsilon (Fig. 5), we used the same trick for these simple LRP configurations, as well. As other LRP rules were already good at disentangling the excitatory and inhibitory regions, we excluded them from this treatment.

### Evaluation of Saliency Maps

We assessed the correspondence between saliency maps and expert annotations via Dice loss [58]: 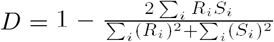, where *R* was a saliency map and *S* the expert annotation. Intuitively, *D ∈* [0, 1] is a normalized distance between *R* and *S*. When a saliency map perfectly matches the expert annotation, *D* decreases to 0. Otherwise, it indicates the degree of mismatch. It is also robust to imbalance between the numbers of foreground and background pixels, which is typically severe due to the relative size of annotations in medical images [58]. However, our post-processing influences *D*. Thus, given a triplet of disease scenario, DNN architecture and attribution method (Fig. 7a,b,d, and e), we searched for the optimal *f*_*ν*_ among 20 values spaced evenly within [0.0005, 1] on a log scale with a geometric^1^ progression. Our criterion was based on the overall (dis)agreement between saliency maps and expert annotations. The optimal values can be found in Table 4 in Appendix A.2. We also show examples of optimally processed saliency maps in Fig. 10 and Fig. 11.

**Table 4:**
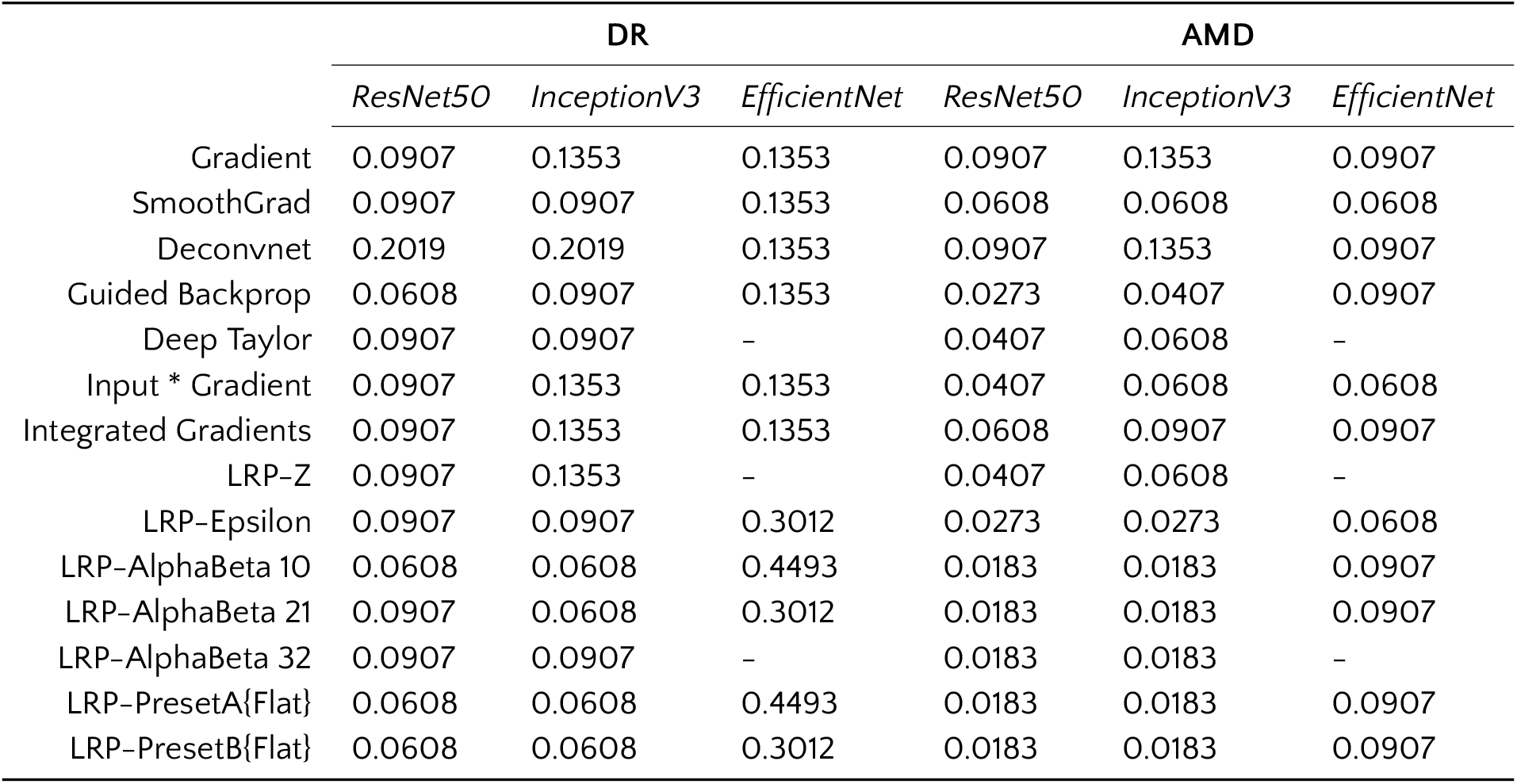
Optimum *f*_*ν*_ values for attribution methods under the DR and AMD scenarios.

|  | DR |  |  | AMD |  |  |
| --- | --- | --- | --- | --- | --- | --- |
|  | *ResNet50* | *InceptionV3* | *EfficientNet* | *ResNet50* | *InceptionV3* | *EfficientNet* |
| Gradient | 0.0907 | 0.1353 | 0.1353 | 0.0907 | 0.1353 | 0.0907 |
| SmoothGrad | 0.0907 | 0.0907 | 0.1353 | 0.0608 | 0.0608 | 0.0608 |
| Deconvnet | 0.2019 | 0.2019 | 0.1353 | 0.0907 | 0.1353 | 0.0907 |
| Guided Backprop | 0.0608 | 0.0907 | 0.1353 | 0.0273 | 0.0407 | 0.0907 |
| Deep Taylor | 0.0907 | 0.0907 | – | 0.0407 | 0.0608 | – |
| Input * Gradient | 0.0907 | 0.1353 | 0.1353 | 0.0407 | 0.0608 | 0.0608 |
| Integrated Gradients | 0.0907 | 0.1353 | 0.1353 | 0.0608 | 0.0907 | 0.0907 |
| LRP-Z | 0.0907 | 0.1353 | – | 0.0407 | 0.0608 | – |
| LRP-Epsilon | 0.0907 | 0.0907 | 0.3012 | 0.0273 | 0.0273 | 0.0608 |
| LRP-AlphaBeta 10 | 0.0608 | 0.0608 | 0.4493 | 0.0183 | 0.0183 | 0.0907 |
| LRP-AlphaBeta 21 | 0.0907 | 0.0608 | 0.3012 | 0.0183 | 0.0183 | 0.0907 |
| LRP-AlphaBeta 32 | 0.0907 | 0.0907 | – | 0.0183 | 0.0183 | – |
| LRP-PresetA{Flat} | 0.0608 | 0.0608 | 0.4493 | 0.0183 | 0.0183 | 0.0907 |
| LRP-PresetB{Flat} | 0.0608 | 0.0608 | 0.3012 | 0.0183 | 0.0183 | 0.0907 |

**Table 5:**
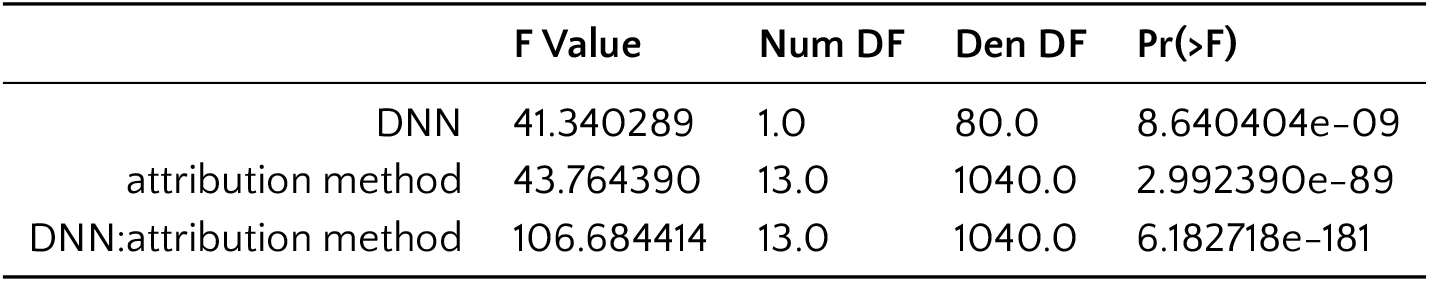
2-way repeated measures ANOVA results, saliency map correspondence to expert annotation via Dice loss. Factor “DNN” can take 2 two values: ResNet50 or InceptionV3; factor “attribution method” can be one of 14 attribution methods: Gradient, SmoothGrad, Deconvnet, Guided Backprop, Deep Taylor, Input * Gradient, Integrated Gradients, LRP-Z, LRP-Epsilon, LRP-AlphaBeta 10, LRP-AlphaBeta 21, LRP-AlphaBeta 32, LRP-PresetA, LRP-PresetB.

|  | F Value | Num DF | Den DF | Pr(>F) |
| --- | --- | --- | --- | --- |
| DNN | 41.340289 | 1.0 | 80.0 | 8.640404e-09 |
| attribution method | 43.764390 | 13.0 | 1040.0 | 2.992390e-89 |
| DNN:attribution method | 106.684414 | 13.0 | 1040.0 | 6.182718e-181 |

**Table 6:** Multiple comparison of attribution methods w.r.t. Dice loss, using Tukey HSD with `alpha=0.05`

| group1 | group2 | meandiff | p-adj | lower | upper | reject |
| --- | --- | --- | --- | --- | --- | --- |
| Guided Backprop | SmoothGrad | 0.009 | 0.9 | -0.0203 | 0.0383 | False |
| Guided Backprop | Integrated Gradients | 0.0593 | 0.001 | 0.0299 | 0.0886 | True |
| Guided Backprop | LRP-Epsilon | 0.0622 | 0.001 | 0.0329 | 0.0915 | True |
| Guided Backprop | LRP-PresetA | 0.0623 | 0.001 | 0.033 | 0.0916 | True |
| Guided Backprop | LRP-PresetB | 0.0623 | 0.001 | 0.033 | 0.0917 | True |
| Guided Backprop | Deep Taylor | 0.0624 | 0.001 | 0.0331 | 0.0917 | True |
| Guided Backprop | LRP-AlphaBeta 10 | 0.0652 | 0.001 | 0.0359 | 0.0945 | True |
| Guided Backprop | LRP-AlphaBeta 21 | 0.0676 | 0.001 | 0.0383 | 0.0969 | True |
| Guided Backprop | Gradient | 0.0683 | 0.001 | 0.039 | 0.0977 | True |
| Guided Backprop | Input * Gradient | 0.0728 | 0.001 | 0.0435 | 0.1022 | True |
| Guided Backprop | LRP-Z | 0.0728 | 0.001 | 0.0435 | 0.1022 | True |
| Guided Backprop | LRP-AlphaBeta 32 | 0.0946 | 0.001 | 0.0652 | 0.1239 | True |
| Guided Backprop | Deconvnet | 0.1276 | 0.001 | 0.0983 | 0.157 | True |
| SmoothGrad | Integrated Gradients | 0.0503 | 0.001 | 0.0209 | 0.0796 | True |
| SmoothGrad | LRP-Epsilon | 0.0532 | 0.001 | 0.0239 | 0.0825 | True |
| SmoothGrad | LRP-PresetA | 0.0533 | 0.001 | 0.024 | 0.0826 | True |
| SmoothGrad | LRP-PresetB | 0.0533 | 0.001 | 0.024 | 0.0827 | True |
| SmoothGrad | Deep Taylor | 0.0534 | 0.001 | 0.0241 | 0.0827 | True |
| SmoothGrad | LRP-AlphaBeta 10 | 0.0562 | 0.001 | 0.0269 | 0.0856 | True |
| SmoothGrad | LRP-AlphaBeta 21 | 0.0586 | 0.001 | 0.0293 | 0.0879 | True |
| SmoothGrad | Gradient | 0.0593 | 0.001 | 0.03 | 0.0887 | True |
| SmoothGrad | Input * Gradient | 0.0638 | 0.001 | 0.0345 | 0.0932 | True |
| SmoothGrad | LRP-Z | 0.0639 | 0.001 | 0.0345 | 0.0932 | True |
| SmoothGrad | LRP-AlphaBeta 32 | 0.0856 | 0.001 | 0.0563 | 0.1149 | True |
| SmoothGrad | Deconvnet | 0.1187 | 0.001 | 0.0893 | 0.148 | True |
| Integrated Gradients | LRP-Epsilon | 0.003 | 0.9 | -0.0264 | 0.0323 | False |
| Integrated Gradients | LRP-PresetA | 0.003 | 0.9 | -0.0263 | 0.0324 | False |
| Integrated Gradients | LRP-PresetB | 0.0031 | 0.9 | -0.0262 | 0.0324 | False |

Table 6 – Continued from previous page
| group1 | group2 | meandiff | p-adj | lower | upper | reject |
| --- | --- | --- | --- | --- | --- | --- |
| Integrated Gradients | Deep Taylor | 0.0031 | 0.9 | -0.0262 | 0.0325 | False |
| Integrated Gradients | LRP-AlphaBeta 10 | 0.006 | 0.9 | -0.0234 | 0.0353 | False |
| Integrated Gradients | LRP-AlphaBeta 21 | 0.0083 | 0.9 | -0.021 | 0.0377 | False |
| Integrated Gradients | Gradient | 0.0091 | 0.9 | -0.0202 | 0.0384 | False |
| Integrated Gradients | Input * Gradient | 0.0136 | 0.9 | -0.0157 | 0.0429 | False |
| Integrated Gradients | LRP-Z | 0.0136 | 0.9 | -0.0157 | 0.0429 | False |
| Integrated Gradients | LRP-AlphaBeta 32 | 0.0353 | 0.0043 | 0.006 | 0.0646 | True |
| Integrated Gradients | Deconvnet | 0.0684 | 0.001 | 0.0391 | 0.0977 | True |
| LRP-Epsilon | LRP-PresetA | 0.0001 | 0.9 | -0.0292 | 0.0294 | False |
| LRP-Epsilon | LRP-PresetB | 0.0001 | 0.9 | -0.0292 | 0.0294 | False |
| LRP-Epsilon | Deep Taylor | 0.0002 | 0.9 | -0.0291 | 0.0295 | False |
| LRP-Epsilon | LRP-AlphaBeta 10 | 0.003 | 0.9 | -0.0263 | 0.0323 | False |
| LRP-Epsilon | LRP-AlphaBeta 21 | 0.0054 | 0.9 | -0.0239 | 0.0347 | False |
| LRP-Epsilon | Gradient | 0.0061 | 0.9 | -0.0232 | 0.0354 | False |
| LRP-Epsilon | Input * Gradient | 0.0106 | 0.9 | -0.0187 | 0.04 | False |
| LRP-Epsilon | LRP-Z | 0.0106 | 0.9 | -0.0187 | 0.04 | False |
| LRP-Epsilon | LRP-AlphaBeta 32 | 0.0324 | 0.0156 | 0.003 | 0.0617 | True |
| LRP-Epsilon | Deconvnet | 0.0654 | 0.001 | 0.0361 | 0.0948 | True |
| LRP-PresetA | LRP-PresetB | 0.0 | 0.9 | -0.0293 | 0.0294 | False |
| LRP-PresetA | Deep Taylor | 0.0001 | 0.9 | -0.0292 | 0.0294 | False |
| LRP-PresetA | LRP-AlphaBeta 10 | 0.0029 | 0.9 | -0.0264 | 0.0323 | False |
| LRP-PresetA | LRP-AlphaBeta 21 | 0.0053 | 0.9 | -0.024 | 0.0346 | False |
| LRP-PresetA | Gradient | 0.006 | 0.9 | -0.0233 | 0.0354 | False |
| LRP-PresetA | Input * Gradient | 0.0105 | 0.9 | -0.0188 | 0.0399 | False |
| LRP-PresetA | LRP-Z | 0.0106 | 0.9 | -0.0188 | 0.0399 | False |
| LRP-PresetA | LRP-AlphaBeta 32 | 0.0323 | 0.0161 | 0.0029 | 0.0616 | True |
| LRP-PresetA | Deconvnet | 0.0653 | 0.001 | 0.036 | 0.0947 | True |
| LRP-PresetB | Deep Taylor | 0.0001 | 0.9 | -0.0293 | 0.0294 | False |
| LRP-PresetB | LRP-AlphaBeta 10 | 0.0029 | 0.9 | -0.0264 | 0.0322 | False |
| LRP-PresetB | LRP-AlphaBeta 21 | 0.0053 | 0.9 | -0.0241 | 0.0346 | False |
| LRP-PresetB | Gradient | 0.006 | 0.9 | -0.0233 | 0.0353 | False |
| LRP-PresetB | Input * Gradient | 0.0105 | 0.9 | -0.0188 | 0.0398 | False |
| LRP-PresetB | LRP-Z | 0.0105 | 0.9 | -0.0188 | 0.0398 | False |
| LRP-PresetB | LRP-AlphaBeta 32 | 0.0322 | 0.0164 | 0.0029 | 0.0616 | True |
| LRP-PresetB | Deconvnet | 0.0653 | 0.001 | 0.036 | 0.0946 | True |
| Deep Taylor | LRP-AlphaBeta 10 | 0.0028 | 0.9 | -0.0265 | 0.0322 | False |
| Deep Taylor | LRP-AlphaBeta 21 | 0.0052 | 0.9 | -0.0241 | 0.0345 | False |
| Deep Taylor | Gradient | 0.0059 | 0.9 | -0.0234 | 0.0353 | False |
| Deep Taylor | Input * Gradient | 0.0104 | 0.9 | -0.0189 | 0.0398 | False |
| Deep Taylor | LRP-Z | 0.0105 | 0.9 | -0.0189 | 0.0398 | False |
| Deep Taylor | LRP-AlphaBeta 32 | 0.0322 | 0.0168 | 0.0028 | 0.0615 | True |
| Deep Taylor | Deconvnet | 0.0652 | 0.001 | 0.0359 | 0.0946 | True |
| LRP-AlphaBeta 10 | LRP-AlphaBeta 21 | 0.0024 | 0.9 | -0.027 | 0.0317 | False |
| LRP-AlphaBeta 10 | Gradient | 0.0031 | 0.9 | -0.0262 | 0.0324 | False |
| LRP-AlphaBeta 10 | Input * Gradient | 0.0076 | 0.9 | -0.0217 | 0.0369 | False |

Table 6 – Continued from previous page
| group1 | group2 | meandiff | p-adj | lower | upper | reject |
| --- | --- | --- | --- | --- | --- | --- |
| LRP-AlphaBeta 10 | LRP-Z | 0.0076 | 0.9 | -0.0217 | 0.0369 | False |
| LRP-AlphaBeta 10 | LRP-AlphaBeta 32 | 0.0293 | 0.0497 | 0.0 | 0.0587 | True |
| LRP-AlphaBeta 10 | Deconvnet | 0.0624 | 0.001 | 0.0331 | 0.0917 | True |
| LRP-AlphaBeta 21 | Gradient | 0.0007 | 0.9 | -0.0286 | 0.0301 | False |
| LRP-AlphaBeta 21 | Input * Gradient | 0.0052 | 0.9 | -0.0241 | 0.0346 | False |
| LRP-AlphaBeta 21 | LRP-Z | 0.0053 | 0.9 | -0.0241 | 0.0346 | False |
| LRP-AlphaBeta 21 | LRP-AlphaBeta 32 | 0.027 | 0.1103 | -0.0024 | 0.0563 | False |
| LRP-AlphaBeta 21 | Deconvnet | 0.06 | 0.001 | 0.0307 | 0.0894 | True |
| Gradient | Input * Gradient | 0.0045 | 0.9 | -0.0248 | 0.0338 | False |
| Gradient | LRP-Z | 0.0045 | 0.9 | -0.0248 | 0.0338 | False |
| Gradient | LRP-AlphaBeta 32 | 0.0262 | 0.1371 | -0.0031 | 0.0556 | False |
| Gradient | Deconvnet | 0.0593 | 0.001 | 0.03 | 0.0886 | True |
| Input * Gradient | LRP-Z | 0.0 | 0.9 | -0.0293 | 0.0293 | False |
| Input * Gradient | LRP-AlphaBeta 32 | 0.0217 | 0.4214 | -0.0076 | 0.0511 | False |
| Input * Gradient | Deconvnet | 0.0548 | 0.001 | 0.0255 | 0.0841 | True |
| LRP-Z | LRP-AlphaBeta 32 | 0.0217 | 0.4219 | -0.0076 | 0.051 | False |
| LRP-Z | Deconvnet | 0.0548 | 0.001 | 0.0255 | 0.0841 | True |
| LRP-AlphaBeta 32 | Deconvnet | 0.0331 | 0.0115 | 0.0037 | 0.0624 | True |

**Table 7:**
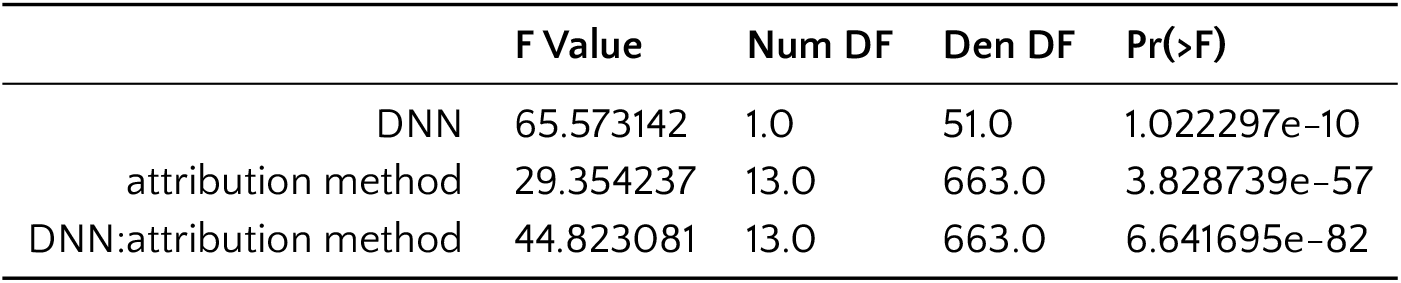
2-way repeated measures ANOVA results, saliency map correspondence to expert annotation via Dice loss. Factor “DNN” can take 2 two values: ResNet50 or InceptionV3; factor “attribution method” can be one of 14 attribution methods: Gradient, SmoothGrad, Deconvnet, Guided Backprop, Deep Taylor, Input * Gradient, Integrated Gradients, LRP-Z, LRP-Epsilon, LRP-AlphaBeta 10, LRP-AlphaBeta 21, LRP-AlphaBeta 32, LRP-PresetA, LRP-PresetB.

|  | F Value | Num DF | Den DF | Pr(>F) |
| --- | --- | --- | --- | --- |
| DNN | 65.573142 | 1.0 | 51.0 | 1.022297e-10 |
| attribution method | 29.354237 | 13.0 | 663.0 | 3.828739e-57 |
| DNN:attribution method | 44.823081 | 13.0 | 663.0 | 6.641695e-82 |

**Table 8:** Multiple comparison of attribution methods w.r.t. Dice loss, using Tukey HSD with `alpha=0.05`

| group1 | group2 | meandiff | p-adj | lower | upper | reject |
| --- | --- | --- | --- | --- | --- | --- |
| Guided Backprop | LRP-PresetBFlat | 0.0138 | 0.9 | -0.0438 | 0.0715 | False |
| Guided Backprop | LRP-PresetAFlat | 0.0273 | 0.9 | -0.0304 | 0.0849 | False |
| Guided Backprop | LRP-AlphaBeta 10 | 0.0386 | 0.5806 | -0.0191 | 0.0963 | False |
| Guided Backprop | LRP-Epsilon | 0.043 | 0.4096 | -0.0147 | 0.1007 | False |
| Guided Backprop | LRP-AlphaBeta 21 | 0.0482 | 0.2231 | -0.0095 | 0.1058 | False |
| Guided Backprop | Deep Taylor | 0.05 | 0.1733 | -0.0077 | 0.1077 | False |
| Guided Backprop | SmoothGrad | 0.0715 | 0.0026 | 0.0138 | 0.1292 | True |

Table 8 – Continued from previous page
| group1 | group2 | meandiff | p-adj | lower | upper | reject |
| --- | --- | --- | --- | --- | --- | --- |
| Guided Backprop | LRP-AlphaBeta 32 | 0.0893 | 0.001 | 0.0316 | 0.147 | True |
| Guided Backprop | LRP-Z | 0.1003 | 0.001 | 0.0427 | 0.158 | True |
| Guided Backprop | Input * Gradient | 0.1003 | 0.001 | 0.0427 | 0.158 | True |
| Guided Backprop | Integrated Gradients | 0.1023 | 0.001 | 0.0446 | 0.16 | True |
| Guided Backprop | Gradient | 0.1555 | 0.001 | 0.0978 | 0.2132 | True |
| Guided Backprop | Deconvnet | 0.2264 | 0.001 | 0.1687 | 0.284 | True |
| LRP-PresetBFlat | LRP-PresetAFlat | 0.0134 | 0.9 | -0.0442 | 0.0711 | False |
| LRP-PresetBFlat | LRP-AlphaBeta 10 | 0.0247 | 0.9 | -0.0329 | 0.0824 | False |
| LRP-PresetBFlat | LRP-Epsilon | 0.0291 | 0.9 | -0.0285 | 0.0868 | False |
| LRP-PresetBFlat | LRP-AlphaBeta 21 | 0.0343 | 0.7391 | -0.0234 | 0.092 | False |
| LRP-PresetBFlat | Deep Taylor | 0.0362 | 0.6702 | -0.0215 | 0.0938 | False |
| LRP-PresetBFlat | SmoothGrad | 0.0577 | 0.0502 | -0.0 | 0.1153 | False |
| LRP-PresetBFlat | LRP-AlphaBeta 32 | 0.0754 | 0.001 | 0.0178 | 0.1331 | True |
| LRP-PresetBFlat | LRP-Z | 0.0865 | 0.001 | 0.0288 | 0.1442 | True |
| LRP-PresetBFlat | Input * Gradient | 0.0865 | 0.001 | 0.0288 | 0.1442 | True |
| LRP-PresetBFlat | Integrated Gradients | 0.0885 | 0.001 | 0.0308 | 0.1461 | True |
| LRP-PresetBFlat | Gradient | 0.1416 | 0.001 | 0.084 | 0.1993 | True |
| LRP-PresetBFlat | Deconvnet | 0.2125 | 0.001 | 0.1549 | 0.2702 | True |
| LRP-PresetAFlat | LRP-AlphaBeta 10 | 0.0113 | 0.9 | -0.0463 | 0.069 | False |
| LRP-PresetAFlat | LRP-Epsilon | 0.0157 | 0.9 | -0.0419 | 0.0734 | False |
| LRP-PresetAFlat | LRP-AlphaBeta 21 | 0.0209 | 0.9 | -0.0368 | 0.0786 | False |
| LRP-PresetAFlat | Deep Taylor | 0.0228 | 0.9 | -0.0349 | 0.0804 | False |
| LRP-PresetAFlat | SmoothGrad | 0.0442 | 0.358 | -0.0134 | 0.1019 | False |
| LRP-PresetAFlat | LRP-AlphaBeta 32 | 0.062 | 0.0217 | 0.0044 | 0.1197 | True |
| LRP-PresetAFlat | LRP-Z | 0.0731 | 0.0018 | 0.0154 | 0.1307 | True |
| LRP-PresetAFlat | Input * Gradient | 0.0731 | 0.0018 | 0.0154 | 0.1307 | True |
| LRP-PresetAFlat | Integrated Gradients | 0.075 | 0.0011 | 0.0174 | 0.1327 | True |
| LRP-PresetAFlat | Gradient | 0.1282 | 0.001 | 0.0706 | 0.1859 | True |
| LRP-PresetAFlat | Deconvnet | 0.1991 | 0.001 | 0.1414 | 0.2568 | True |
| LRP-AlphaBeta 10 | LRP-Epsilon | 0.0044 | 0.9 | -0.0533 | 0.0621 | False |
| LRP-AlphaBeta 10 | LRP-AlphaBeta 21 | 0.0096 | 0.9 | -0.0481 | 0.0672 | False |
| LRP-AlphaBeta 10 | Deep Taylor | 0.0114 | 0.9 | -0.0462 | 0.0691 | False |
| LRP-AlphaBeta 10 | SmoothGrad | 0.0329 | 0.7912 | -0.0248 | 0.0906 | False |
| LRP-AlphaBeta 10 | LRP-AlphaBeta 32 | 0.0507 | 0.1568 | -0.007 | 0.1084 | False |
| LRP-AlphaBeta 10 | LRP-Z | 0.0617 | 0.0229 | 0.0041 | 0.1194 | True |
| LRP-AlphaBeta 10 | Input * Gradient | 0.0617 | 0.0229 | 0.0041 | 0.1194 | True |
| LRP-AlphaBeta 10 | Integrated Gradients | 0.0637 | 0.0153 | 0.006 | 0.1214 | True |
| LRP-AlphaBeta 10 | Gradient | 0.1169 | 0.001 | 0.0592 | 0.1746 | True |
| LRP-AlphaBeta 10 | Deconvnet | 0.1878 | 0.001 | 0.1301 | 0.2455 | True |
| LRP-Epsilon | LRP-AlphaBeta 21 | 0.0052 | 0.9 | -0.0525 | 0.0628 | False |
| LRP-Epsilon | Deep Taylor | 0.007 | 0.9 | -0.0506 | 0.0647 | False |
| LRP-Epsilon | SmoothGrad | 0.0285 | 0.9 | -0.0292 | 0.0862 | False |
| LRP-Epsilon | LRP-AlphaBeta 32 | 0.0463 | 0.282 | -0.0114 | 0.104 | False |
| LRP-Epsilon | LRP-Z | 0.0573 | 0.0531 | -0.0003 | 0.115 | False |
| LRP-Epsilon | Input * Gradient | 0.0573 | 0.0531 | -0.0003 | 0.115 | False |

Table 8 – *Continued from previous page*
| group1 | group2 | meandiff | p-adj | lower | upper | reject |
| --- | --- | --- | --- | --- | --- | --- |
| LRP-Epsilon | Integrated Gradients | 0.0593 | 0.037 | 0.0016 | 0.117 | True |
| LRP-Epsilon | Gradient | 0.1125 | 0.001 | 0.0548 | 0.1702 | True |
| LRP-Epsilon | Deconvnet | 0.1834 | 0.001 | 0.1257 | 0.2411 | True |
| LRP-AlphaBeta 21 | Deep Taylor | 0.0019 | 0.9 | -0.0558 | 0.0595 | False |
| LRP-AlphaBeta 21 | SmoothGrad | 0.0233 | 0.9 | -0.0343 | 0.081 | False |
| LRP-AlphaBeta 21 | LRP-AlphaBeta 32 | 0.0411 | 0.4856 | -0.0165 | 0.0988 | False |
| LRP-AlphaBeta 21 | LRP-Z | 0.0522 | 0.1251 | -0.0055 | 0.1098 | False |
| LRP-AlphaBeta 21 | Input * Gradient | 0.0522 | 0.125 | -0.0055 | 0.1098 | False |
| LRP-AlphaBeta 21 | Integrated Gradients | 0.0541 | 0.0917 | -0.0035 | 0.1118 | False |
| LRP-AlphaBeta 21 | Gradient | 0.1073 | 0.001 | 0.0497 | 0.165 | True |
| LRP-AlphaBeta 21 | Deconvnet | 0.1782 | 0.001 | 0.1206 | 0.2359 | True |
| Deep Taylor | SmoothGrad | 0.0215 | 0.9 | -0.0362 | 0.0791 | False |
| Deep Taylor | LRP-AlphaBeta 32 | 0.0393 | 0.5551 | -0.0184 | 0.0969 | False |
| Deep Taylor | LRP-Z | 0.0503 | 0.1659 | -0.0074 | 0.108 | False |
| Deep Taylor | Input * Gradient | 0.0503 | 0.1659 | -0.0074 | 0.108 | False |
| Deep Taylor | Integrated Gradients | 0.0523 | 0.1243 | -0.0054 | 0.11 | False |
| Deep Taylor | Gradient | 0.1055 | 0.001 | 0.0478 | 0.1631 | True |
| Deep Taylor | Deconvnet | 0.1764 | 0.001 | 0.1187 | 0.234 | True |
| SmoothGrad | LRP-AlphaBeta 32 | 0.0178 | 0.9 | -0.0399 | 0.0755 | False |
| SmoothGrad | LRP-Z | 0.0288 | 0.9 | -0.0288 | 0.0865 | False |
| SmoothGrad | Input * Gradient | 0.0288 | 0.9 | -0.0288 | 0.0865 | False |
| SmoothGrad | Integrated Gradients | 0.0308 | 0.8695 | -0.0269 | 0.0885 | False |
| SmoothGrad | Gradient | 0.084 | 0.001 | 0.0263 | 0.1417 | True |
| SmoothGrad | Deconvnet | 0.1549 | 0.001 | 0.0972 | 0.2126 | True |
| LRP-AlphaBeta 32 | LRP-Z | 0.011 | 0.9 | -0.0466 | 0.0687 | False |
| LRP-AlphaBeta 32 | Input * Gradient | 0.011 | 0.9 | -0.0466 | 0.0687 | False |
| LRP-AlphaBeta 32 | Integrated Gradients | 0.013 | 0.9 | -0.0447 | 0.0707 | False |
| LRP-AlphaBeta 32 | Gradient | 0.0662 | 0.009 | 0.0085 | 0.1239 | True |
| LRP-AlphaBeta 32 | Deconvnet | 0.1371 | 0.001 | 0.0794 | 0.1948 | True |
| LRP-Z | Input * Gradient | 0.0 | 0.9 | -0.0577 | 0.0577 | False |
| LRP-Z | Integrated Gradients | 0.002 | 0.9 | -0.0557 | 0.0596 | False |
| LRP-Z | Gradient | 0.0552 | 0.0782 | -0.0025 | 0.1128 | False |
| LRP-Z | Deconvnet | 0.1261 | 0.001 | 0.0684 | 0.1837 | True |
| Input * Gradient | Integrated Gradients | 0.002 | 0.9 | -0.0557 | 0.0596 | False |
| Input * Gradient | Gradient | 0.0552 | 0.0782 | -0.0025 | 0.1128 | False |
| Input * Gradient | Deconvnet | 0.126 | 0.001 | 0.0684 | 0.1837 | True |
| Integrated Gradients | Gradient | 0.0532 | 0.1074 | -0.0045 | 0.1109 | False |
| Integrated Gradients | Deconvnet | 0.1241 | 0.001 | 0.0664 | 0.1818 | True |
| Gradient | Deconvnet | 0.0709 | 0.0031 | 0.0132 | 0.1286 | True |

**Table 9:** 2-way repeated measures ANOVA results, average relative softmax difference. Factor “DNN” can take 2 two values: ResNet50 or InceptionV3; factor “attribution method” can be one of 16 attribution methods: Gradient, SmoothGrad, Deconvnet, Guided Backprop, Deep Taylor, Input * Gradient, Integrated Gradients, LRP-Z, LRP-Epsilon, LRP-AlphaBeta 10, LRP-AlphaBeta 21, LRP-AlphaBeta 32, LRP-PresetA, LRP-PresetB, Random, Expert

|  | F Value | Num DF | Den DF | Pr(>F) |
| --- | --- | --- | --- | --- |
| DNN | 1.900759 | 1.0 | 80.0 | 1.718364e-01 |
| attribution method | 113.691369 | 15.0 | 1200.0 | 7.772892e-218 |
| DNN:attribution method | 5.465855 | 15.0 | 1200.0 | 7.808541e-11 |

**Table 10:** Multiple comparison of attribution methodsw.r.t. average relative softmax difference, using Tukey HSD with `alpha=0.05`

| group1 | group2 | meandiff | p-adj | lower | upper | reject |
| --- | --- | --- | --- | --- | --- | --- |
| Guided Backprop | LRP-PresetB | 0.0383 | 0.9 | -0.056 | 0.1326 | False |
| Guided Backprop | LRP-AlphaBeta 21 | 0.0486 | 0.9 | -0.0457 | 0.1428 | False |
| Guided Backprop | Expert | 0.0647 | 0.5701 | -0.0296 | 0.159 | False |
| Guided Backprop | Integrated Gradients | 0.0677 | 0.5005 | -0.0266 | 0.162 | False |
| Guided Backprop | LRP-Epsilon | 0.0706 | 0.4276 | -0.0237 | 0.1649 | False |
| Guided Backprop | LRP-AlphaBeta 32 | 0.0759 | 0.2936 | -0.0184 | 0.1702 | False |
| Guided Backprop | LRP-PresetA | 0.0815 | 0.1847 | -0.0128 | 0.1758 | False |
| Guided Backprop | LRP-AlphaBeta 10 | 0.101 | 0.0222 | 0.0067 | 0.1953 | True |
| Guided Backprop | Gradient | 0.1041 | 0.0148 | 0.0098 | 0.1983 | True |
| Guided Backprop | LRP-Z | 0.1236 | 0.001 | 0.0293 | 0.2179 | True |
| Guided Backprop | Input * Gradient | 0.1237 | 0.001 | 0.0294 | 0.218 | True |
| Guided Backprop | Deep Taylor | 0.1248 | 0.001 | 0.0305 | 0.219 | True |
| Guided Backprop | Deconvnet | 0.3301 | 0.001 | 0.2358 | 0.4244 | True |
| Guided Backprop | SmoothGrad | 0.381 | 0.001 | 0.2867 | 0.4753 | True |
| Guided Backprop | Random | 0.5006 | 0.001 | 0.4063 | 0.5949 | True |
| LRP-PresetB | LRP-AlphaBeta 21 | 0.0103 | 0.9 | -0.084 | 0.1046 | False |
| LRP-PresetB | Expert | 0.0265 | 0.9 | -0.0678 | 0.1208 | False |
| LRP-PresetB | Integrated Gradients | 0.0294 | 0.9 | -0.0649 | 0.1237 | False |
| LRP-PresetB | LRP-Epsilon | 0.0323 | 0.9 | -0.062 | 0.1266 | False |
| LRP-PresetB | LRP-AlphaBeta 32 | 0.0376 | 0.9 | -0.0567 | 0.1319 | False |
| LRP-PresetB | LRP-PresetA | 0.0432 | 0.9 | -0.0511 | 0.1375 | False |
| LRP-PresetB | LRP-AlphaBeta 10 | 0.0627 | 0.6179 | -0.0316 | 0.157 | False |
| LRP-PresetB | Gradient | 0.0658 | 0.5453 | -0.0285 | 0.1601 | False |
| LRP-PresetB | LRP-Z | 0.0853 | 0.1288 | -0.0089 | 0.1796 | False |
| LRP-PresetB | Input * Gradient | 0.0854 | 0.128 | -0.0089 | 0.1797 | False |
| LRP-PresetB | Deep Taylor | 0.0865 | 0.1163 | -0.0078 | 0.1808 | False |
| LRP-PresetB | Deconvnet | 0.2918 | 0.001 | 0.1975 | 0.3861 | True |
| LRP-PresetB | SmoothGrad | 0.3427 | 0.001 | 0.2484 | 0.437 | True |
| LRP-PresetB | Random | 0.4623 | 0.001 | 0.3681 | 0.5566 | True |
| LRP-AlphaBeta 21 | Expert | 0.0162 | 0.9 | -0.0781 | 0.1105 | False |

Table 10 – *Continued from previous page*
| group1 | group2 | meandiff | p-adj | lower | upper | reject |
| --- | --- | --- | --- | --- | --- | --- |
| LRP-AlphaBeta 21 | Integrated Gradients | 0.0191 | 0.9 | -0.0752 | 0.1134 | False |
| LRP-AlphaBeta 21 | LRP-Epsilon | 0.022 | 0.9 | -0.0723 | 0.1163 | False |
| LRP-AlphaBeta 21 | LRP-AlphaBeta 32 | 0.0273 | 0.9 | -0.067 | 0.1216 | False |
| LRP-AlphaBeta 21 | LRP-PresetA | 0.0329 | 0.9 | -0.0613 | 0.1272 | False |
| LRP-AlphaBeta 21 | LRP-AlphaBeta 10 | 0.0524 | 0.8599 | -0.0419 | 0.1467 | False |
| LRP-AlphaBeta 21 | Gradient | 0.0555 | 0.7873 | -0.0388 | 0.1498 | False |
| LRP-AlphaBeta 21 | LRP-Z | 0.0751 | 0.3121 | -0.0192 | 0.1693 | False |
| LRP-AlphaBeta 21 | Input * Gradient | 0.0751 | 0.3107 | -0.0192 | 0.1694 | False |
| LRP-AlphaBeta 21 | Deep Taylor | 0.0762 | 0.2868 | -0.0181 | 0.1705 | False |
| LRP-AlphaBeta 21 | Deconvnet | 0.2815 | 0.001 | 0.1872 | 0.3758 | True |
| LRP-AlphaBeta 21 | SmoothGrad | 0.3324 | 0.001 | 0.2381 | 0.4267 | True |
| LRP-AlphaBeta 21 | Random | 0.4521 | 0.001 | 0.3578 | 0.5463 | True |
| Expert | Integrated Gradients | 0.003 | 0.9 | -0.0913 | 0.0973 | False |
| Expert | LRP-Epsilon | 0.0058 | 0.9 | -0.0885 | 0.1001 | False |
| Expert | LRP-AlphaBeta 32 | 0.0112 | 0.9 | -0.0831 | 0.1054 | False |
| Expert | LRP-PresetA | 0.0168 | 0.9 | -0.0775 | 0.1111 | False |
| Expert | LRP-AlphaBeta 10 | 0.0362 | 0.9 | -0.0581 | 0.1305 | False |
| Expert | Gradient | 0.0393 | 0.9 | -0.055 | 0.1336 | False |
| Expert | LRP-Z | 0.0589 | 0.7076 | -0.0354 | 0.1532 | False |
| Expert | Input * Gradient | 0.0589 | 0.7061 | -0.0353 | 0.1532 | False |
| Expert | Deep Taylor | 0.06 | 0.6809 | -0.0343 | 0.1543 | False |
| Expert | Deconvnet | 0.2653 | 0.001 | 0.1711 | 0.3596 | True |
| Expert | SmoothGrad | 0.3163 | 0.001 | 0.222 | 0.4105 | True |
| Expert | Random | 0.4359 | 0.001 | 0.3416 | 0.5302 | True |
| Integrated Gradients | LRP-Epsilon | 0.0029 | 0.9 | -0.0914 | 0.0972 | False |
| Integrated Gradients | LRP-AlphaBeta 32 | 0.0082 | 0.9 | -0.0861 | 0.1025 | False |
| Integrated Gradients | LRP-PresetA | 0.0138 | 0.9 | -0.0805 | 0.1081 | False |
| Integrated Gradients | LRP-AlphaBeta 10 | 0.0333 | 0.9 | -0.061 | 0.1276 | False |
| Integrated Gradients | Gradient | 0.0364 | 0.9 | -0.0579 | 0.1307 | False |
| Integrated Gradients | LRP-Z | 0.0559 | 0.7773 | -0.0384 | 0.1502 | False |
| Integrated Gradients | Input * Gradient | 0.056 | 0.7758 | -0.0383 | 0.1503 | False |
| Integrated Gradients | Deep Taylor | 0.0571 | 0.7506 | -0.0372 | 0.1513 | False |
| Integrated Gradients | Deconvnet | 0.2624 | 0.001 | 0.1681 | 0.3567 | True |
| Integrated Gradients | SmoothGrad | 0.3133 | 0.001 | 0.219 | 0.4076 | True |
| Integrated Gradients | Random | 0.4329 | 0.001 | 0.3386 | 0.5272 | True |
| LRP-Epsilon | LRP-AlphaBeta 32 | 0.0053 | 0.9 | -0.089 | 0.0996 | False |
| LRP-Epsilon | LRP-PresetA | 0.0109 | 0.9 | -0.0833 | 0.1052 | False |
| LRP-Epsilon | LRP-AlphaBeta 10 | 0.0304 | 0.9 | -0.0639 | 0.1247 | False |
| LRP-Epsilon | Gradient | 0.0335 | 0.9 | -0.0608 | 0.1278 | False |
| LRP-Epsilon | LRP-Z | 0.0531 | 0.8448 | -0.0412 | 0.1473 | False |
| LRP-Epsilon | Input * Gradient | 0.0531 | 0.8433 | -0.0412 | 0.1474 | False |
| LRP-Epsilon | Deep Taylor | 0.0542 | 0.818 | -0.0401 | 0.1485 | False |
| LRP-Epsilon | Deconvnet | 0.2595 | 0.001 | 0.1652 | 0.3538 | True |
| LRP-Epsilon | SmoothGrad | 0.3104 | 0.001 | 0.2161 | 0.4047 | True |
| LRP-Epsilon | Random | 0.43 | 0.001 | 0.3358 | 0.5243 | True |

Table 10 – Continued from previous page
| group1 | group2 | meandiff | p-adj | lower | upper | reject |
| --- | --- | --- | --- | --- | --- | --- |
| LRP-AlphaBeta 32 | LRP-PresetA | 0.0056 | 0.9 | -0.0887 | 0.0999 | False |
| LRP-AlphaBeta 32 | LRP-AlphaBeta 10 | 0.0251 | 0.9 | -0.0692 | 0.1194 | False |
| LRP-AlphaBeta 32 | Gradient | 0.0282 | 0.9 | -0.0661 | 0.1225 | False |
| LRP-AlphaBeta 32 | LRP-Z | 0.0477 | 0.9 | -0.0466 | 0.142 | False |
| LRP-AlphaBeta 32 | Input * Gradient | 0.0478 | 0.9 | -0.0465 | 0.1421 | False |
| LRP-AlphaBeta 32 | Deep Taylor | 0.0489 | 0.9 | -0.0454 | 0.1432 | False |
| LRP-AlphaBeta 32 | Deconvnet | 0.2542 | 0.001 | 0.1599 | 0.3485 | True |
| LRP-AlphaBeta 32 | SmoothGrad | 0.3051 | 0.001 | 0.2108 | 0.3994 | True |
| LRP-AlphaBeta 32 | Random | 0.4247 | 0.001 | 0.3304 | 0.519 | True |
| LRP-PresetA | LRP-AlphaBeta 10 | 0.0195 | 0.9 | -0.0748 | 0.1138 | False |
| LRP-PresetA | Gradient | 0.0226 | 0.9 | -0.0717 | 0.1168 | False |
| LRP-PresetA | LRP-Z | 0.0421 | 0.9 | -0.0522 | 0.1364 | False |
| LRP-PresetA | Input * Gradient | 0.0422 | 0.9 | -0.0521 | 0.1365 | False |
| LRP-PresetA | Deep Taylor | 0.0433 | 0.9 | -0.051 | 0.1375 | False |
| LRP-PresetA | Deconvnet | 0.2486 | 0.001 | 0.1543 | 0.3429 | True |
| LRP-PresetA | SmoothGrad | 0.2995 | 0.001 | 0.2052 | 0.3938 | True |
| LRP-PresetA | Random | 0.4191 | 0.001 | 0.3248 | 0.5134 | True |
| LRP-AlphaBeta 10 | Gradient | 0.0031 | 0.9 | -0.0912 | 0.0974 | False |
| LRP-AlphaBeta 10 | LRP-Z | 0.0226 | 0.9 | -0.0716 | 0.1169 | False |
| LRP-AlphaBeta 10 | Input * Gradient | 0.0227 | 0.9 | -0.0716 | 0.117 | False |
| LRP-AlphaBeta 10 | Deep Taylor | 0.0238 | 0.9 | -0.0705 | 0.1181 | False |
| LRP-AlphaBeta 10 | Deconvnet | 0.2291 | 0.001 | 0.1348 | 0.3234 | True |
| LRP-AlphaBeta 10 | SmoothGrad | 0.28 | 0.001 | 0.1857 | 0.3743 | True |
| LRP-AlphaBeta 10 | Random | 0.3996 | 0.001 | 0.3054 | 0.4939 | True |
| Gradient | LRP-Z | 0.0196 | 0.9 | -0.0747 | 0.1138 | False |
| Gradient | Input * Gradient | 0.0196 | 0.9 | -0.0747 | 0.1139 | False |
| Gradient | Deep Taylor | 0.0207 | 0.9 | -0.0736 | 0.115 | False |
| Gradient | Deconvnet | 0.226 | 0.001 | 0.1317 | 0.3203 | True |
| Gradient | SmoothGrad | 0.2769 | 0.001 | 0.1826 | 0.3712 | True |
| Gradient | Random | 0.3966 | 0.001 | 0.3023 | 0.4908 | True |
| LRP-Z | Input * Gradient | 0.0001 | 0.9 | -0.0942 | 0.0944 | False |
| LRP-Z | Deep Taylor | 0.0011 | 0.9 | -0.0932 | 0.0954 | False |
| LRP-Z | Deconvnet | 0.2065 | 0.001 | 0.1122 | 0.3007 | True |
| LRP-Z | SmoothGrad | 0.2574 | 0.001 | 0.1631 | 0.3517 | True |
| LRP-Z | Random | 0.377 | 0.001 | 0.2827 | 0.4713 | True |
| Input * Gradient | Deep Taylor | 0.0011 | 0.9 | -0.0932 | 0.0954 | False |
| Input * Gradient | Deconvnet | 0.2064 | 0.001 | 0.1121 | 0.3007 | True |
| Input * Gradient | SmoothGrad | 0.2573 | 0.001 | 0.163 | 0.3516 | True |
| Input * Gradient | Random | 0.3769 | 0.001 | 0.2826 | 0.4712 | True |
| Deep Taylor | Deconvnet | 0.2053 | 0.001 | 0.111 | 0.2996 | True |
| Deep Taylor | SmoothGrad | 0.2562 | 0.001 | 0.1619 | 0.3505 | True |
| Deep Taylor | Random | 0.3759 | 0.001 | 0.2816 | 0.4701 | True |
| Deconvnet | SmoothGrad | 0.0509 | 0.8951 | -0.0434 | 0.1452 | False |
| Deconvnet | Random | 0.1705 | 0.001 | 0.0763 | 0.2648 | True |
| SmoothGrad | Random | 0.1196 | 0.0015 | 0.0253 | 0.2139 | True |

**Table 11:** 2-way repeated measures ANOVA results, average relative softmax difference. Factor “DNN” can take 2 two values: ResNet50 or InceptionV3; factor “attribution method” can be one of 16 attribution methods: Gradient, SmoothGrad, Deconvnet, Guided Backprop, Deep Taylor, Input * Gradient, Integrated Gradients, LRP-Z, LRP-Epsilon, LRP-AlphaBeta 10, LRP-AlphaBeta 21, LRP-AlphaBeta 32, LRP-PresetA, LRP-PresetB, Random, Expert

|  | F Value | Num DF | Den DF | Pr(>F) |
| --- | --- | --- | --- | --- |
| DNN | 0.189160 | 1.0 | 51.0 | 6.654512e-01 |
| attribution method | 116.869091 | 15.0 | 765.0 | 4.146892e-186 |
| DNN:attribution method | 6.004318 | 15.0 | 765.0 | 5.830063e-12 |

**Table 12:**
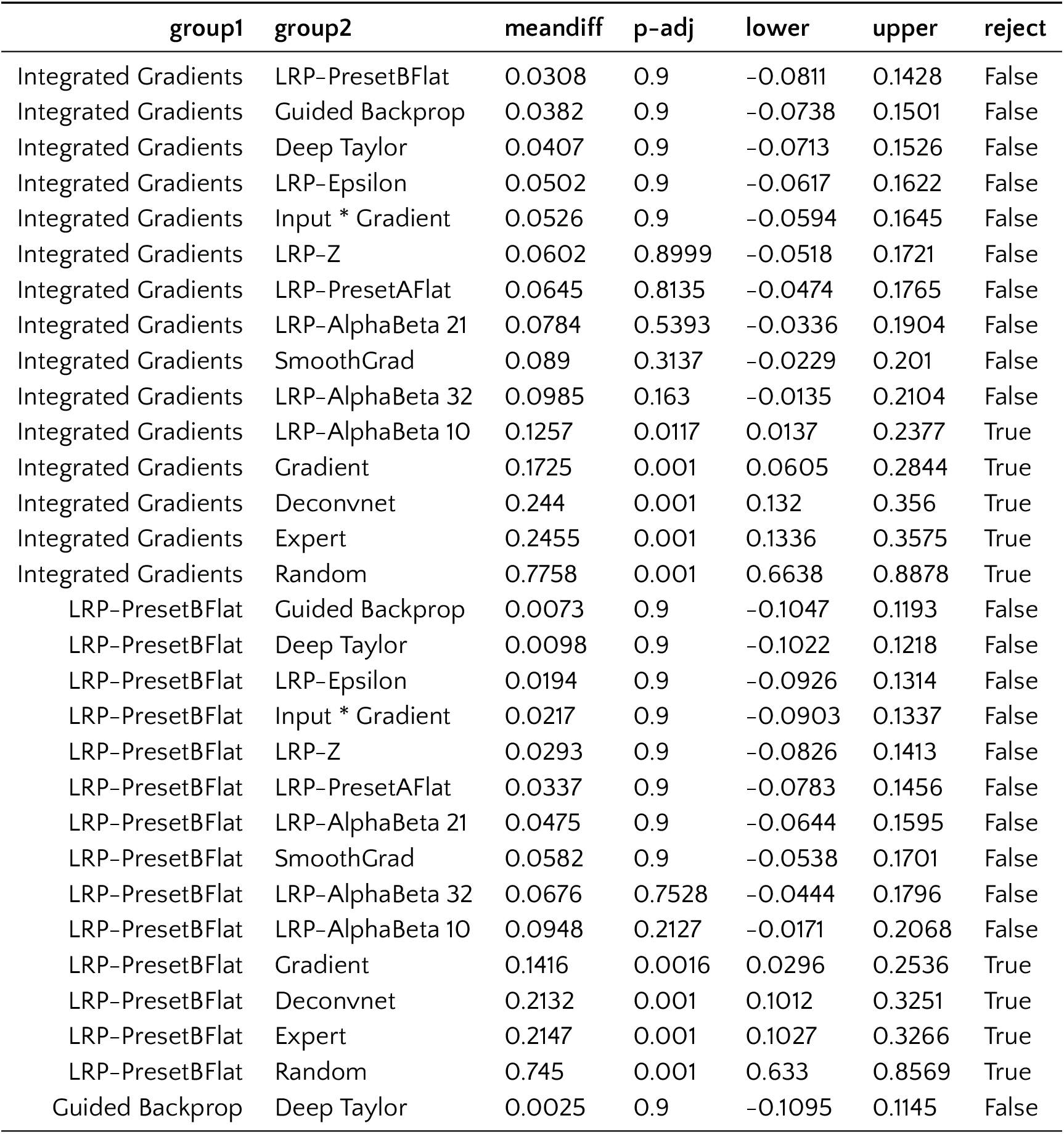
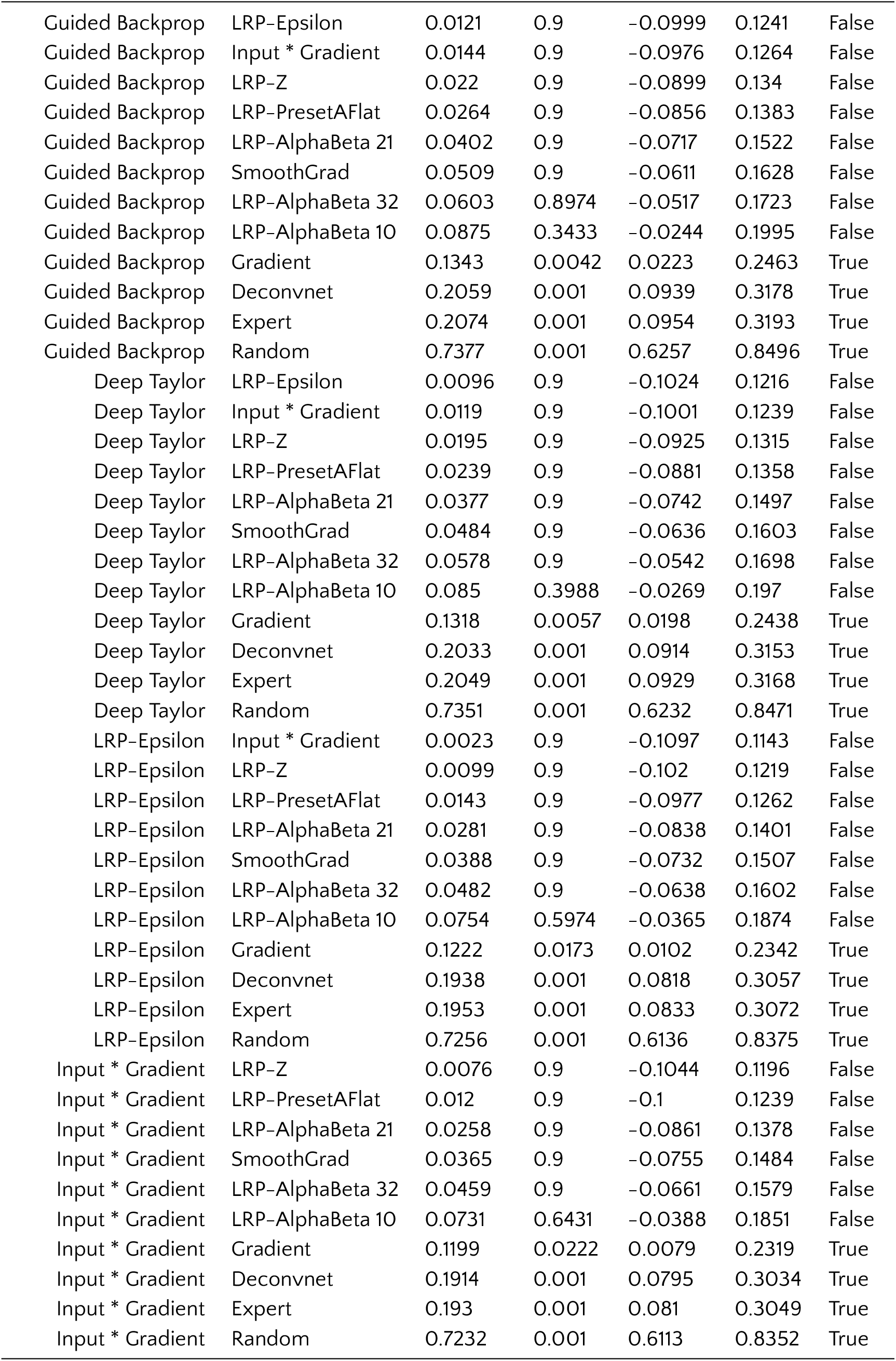
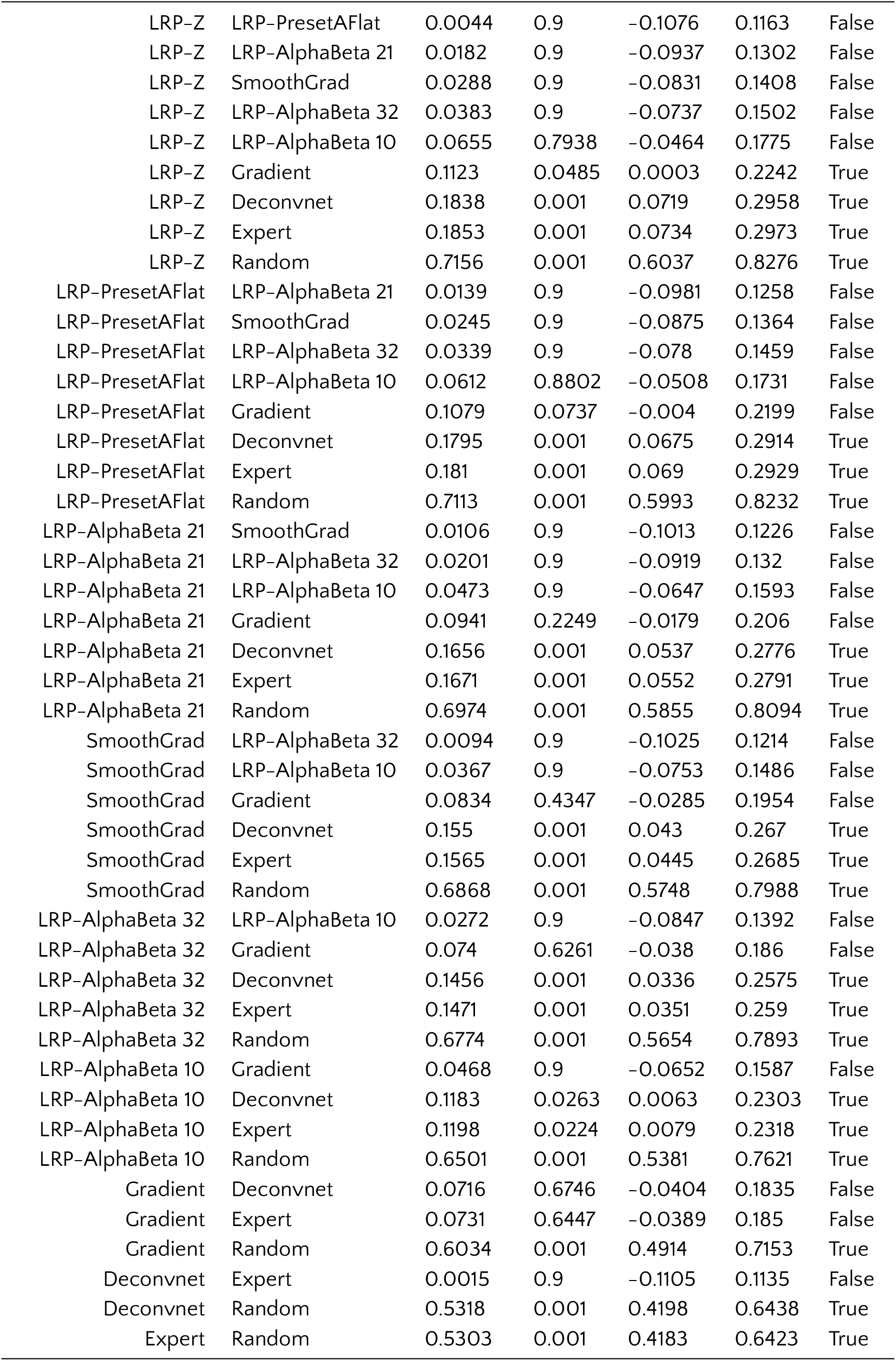
Multiple comparison of attribution methods w.r.t. average relative softmax difference, using Tukey HSD with `alpha=0.05`

| group1 | group2 | meandiff | p-adj | lower | upper | reject |
| --- | --- | --- | --- | --- | --- | --- |
| Integrated Gradients | LRP-PresetBFlat | 0.0308 | 0.9 | -0.0811 | 0.1428 | False |
| Integrated Gradients | Guided Backprop | 0.0382 | 0.9 | -0.0738 | 0.1501 | False |
| Integrated Gradients | Deep Taylor | 0.0407 | 0.9 | -0.0713 | 0.1526 | False |
| Integrated Gradients | LRP-Epsilon | 0.0502 | 0.9 | -0.0617 | 0.1622 | False |
| Integrated Gradients | Input * Gradient | 0.0526 | 0.9 | -0.0594 | 0.1645 | False |
| Integrated Gradients | LRP-Z | 0.0602 | 0.8999 | -0.0518 | 0.1721 | False |
| Integrated Gradients | LRP-PresetAFlat | 0.0645 | 0.8135 | -0.0474 | 0.1765 | False |
| Integrated Gradients | LRP-AlphaBeta 21 | 0.0784 | 0.5393 | -0.0336 | 0.1904 | False |
| Integrated Gradients | SmoothGrad | 0.089 | 0.3137 | -0.0229 | 0.201 | False |
| Integrated Gradients | LRP-AlphaBeta 32 | 0.0985 | 0.163 | -0.0135 | 0.2104 | False |
| Integrated Gradients | LRP-AlphaBeta 10 | 0.1257 | 0.0117 | 0.0137 | 0.2377 | True |
| Integrated Gradients | Gradient | 0.1725 | 0.001 | 0.0605 | 0.2844 | True |
| Integrated Gradients | Deconvnet | 0.244 | 0.001 | 0.132 | 0.356 | True |
| Integrated Gradients | Expert | 0.2455 | 0.001 | 0.1336 | 0.3575 | True |
| Integrated Gradients | Random | 0.7758 | 0.001 | 0.6638 | 0.8878 | True |
| LRP-PresetBFlat | Guided Backprop | 0.0073 | 0.9 | -0.1047 | 0.1193 | False |
| LRP-PresetBFlat | Deep Taylor | 0.0098 | 0.9 | -0.1022 | 0.1218 | False |
| LRP-PresetBFlat | LRP-Epsilon | 0.0194 | 0.9 | -0.0926 | 0.1314 | False |
| LRP-PresetBFlat | Input * Gradient | 0.0217 | 0.9 | -0.0903 | 0.1337 | False |
| LRP-PresetBFlat | LRP-Z | 0.0293 | 0.9 | -0.0826 | 0.1413 | False |
| LRP-PresetBFlat | LRP-PresetAFlat | 0.0337 | 0.9 | -0.0783 | 0.1456 | False |
| LRP-PresetBFlat | LRP-AlphaBeta 21 | 0.0475 | 0.9 | -0.0644 | 0.1595 | False |
| LRP-PresetBFlat | SmoothGrad | 0.0582 | 0.9 | -0.0538 | 0.1701 | False |
| LRP-PresetBFlat | LRP-AlphaBeta 32 | 0.0676 | 0.7528 | -0.0444 | 0.1796 | False |
| LRP-PresetBFlat | LRP-AlphaBeta 10 | 0.0948 | 0.2127 | -0.0171 | 0.2068 | False |
| LRP-PresetBFlat | Gradient | 0.1416 | 0.0016 | 0.0296 | 0.2536 | True |
| LRP-PresetBFlat | Deconvnet | 0.2132 | 0.001 | 0.1012 | 0.3251 | True |
| LRP-PresetBFlat | Expert | 0.2147 | 0.001 | 0.1027 | 0.3266 | True |
| LRP-PresetBFlat | Random | 0.745 | 0.001 | 0.633 | 0.8569 | True |
| Guided Backprop | Deep Taylor | 0.0025 | 0.9 | -0.1095 | 0.1145 | False |

Table 12 – *Continued from previous page*
| group1 | group2 | meandiff | p-adj | lower | upper | reject |
| --- | --- | --- | --- | --- | --- | --- |
| Guided Backprop | LRP-Epsilon | 0.0121 | 0.9 | -0.0999 | 0.1241 | False |
| Guided Backprop | Input * Gradient | 0.0144 | 0.9 | -0.0976 | 0.1264 | False |
| Guided Backprop | LRP-Z | 0.022 | 0.9 | -0.0899 | 0.134 | False |
| Guided Backprop | LRP-PresetAFlat | 0.0264 | 0.9 | -0.0856 | 0.1383 | False |
| Guided Backprop | LRP-AlphaBeta 21 | 0.0402 | 0.9 | -0.0717 | 0.1522 | False |
| Guided Backprop | SmoothGrad | 0.0509 | 0.9 | -0.0611 | 0.1628 | False |
| Guided Backprop | LRP-AlphaBeta 32 | 0.0603 | 0.8974 | -0.0517 | 0.1723 | False |
| Guided Backprop | LRP-AlphaBeta 10 | 0.0875 | 0.3433 | -0.0244 | 0.1995 | False |
| Guided Backprop | Gradient | 0.1343 | 0.0042 | 0.0223 | 0.2463 | True |
| Guided Backprop | Deconvnet | 0.2059 | 0.001 | 0.0939 | 0.3178 | True |
| Guided Backprop | Expert | 0.2074 | 0.001 | 0.0954 | 0.3193 | True |
| Guided Backprop | Random | 0.7377 | 0.001 | 0.6257 | 0.8496 | True |
| Deep Taylor | LRP-Epsilon | 0.0096 | 0.9 | -0.1024 | 0.1216 | False |
| Deep Taylor | Input * Gradient | 0.0119 | 0.9 | -0.1001 | 0.1239 | False |
| Deep Taylor | LRP-Z | 0.0195 | 0.9 | -0.0925 | 0.1315 | False |
| Deep Taylor | LRP-PresetAFlat | 0.0239 | 0.9 | -0.0881 | 0.1358 | False |
| Deep Taylor | LRP-AlphaBeta 21 | 0.0377 | 0.9 | -0.0742 | 0.1497 | False |
| Deep Taylor | SmoothGrad | 0.0484 | 0.9 | -0.0636 | 0.1603 | False |
| Deep Taylor | LRP-AlphaBeta 32 | 0.0578 | 0.9 | -0.0542 | 0.1698 | False |
| Deep Taylor | LRP-AlphaBeta 10 | 0.085 | 0.3988 | -0.0269 | 0.197 | False |
| Deep Taylor | Gradient | 0.1318 | 0.0057 | 0.0198 | 0.2438 | True |
| Deep Taylor | Deconvnet | 0.2033 | 0.001 | 0.0914 | 0.3153 | True |
| Deep Taylor | Expert | 0.2049 | 0.001 | 0.0929 | 0.3168 | True |
| Deep Taylor | Random | 0.7351 | 0.001 | 0.6232 | 0.8471 | True |
| LRP-Epsilon | Input * Gradient | 0.0023 | 0.9 | -0.1097 | 0.1143 | False |
| LRP-Epsilon | LRP-Z | 0.0099 | 0.9 | -0.102 | 0.1219 | False |
| LRP-Epsilon | LRP-PresetAFlat | 0.0143 | 0.9 | -0.0977 | 0.1262 | False |
| LRP-Epsilon | LRP-AlphaBeta 21 | 0.0281 | 0.9 | -0.0838 | 0.1401 | False |
| LRP-Epsilon | SmoothGrad | 0.0388 | 0.9 | -0.0732 | 0.1507 | False |
| LRP-Epsilon | LRP-AlphaBeta 32 | 0.0482 | 0.9 | -0.0638 | 0.1602 | False |
| LRP-Epsilon | LRP-AlphaBeta 10 | 0.0754 | 0.5974 | -0.0365 | 0.1874 | False |
| LRP-Epsilon | Gradient | 0.1222 | 0.0173 | 0.0102 | 0.2342 | True |
| LRP-Epsilon | Deconvnet | 0.1938 | 0.001 | 0.0818 | 0.3057 | True |
| LRP-Epsilon | Expert | 0.1953 | 0.001 | 0.0833 | 0.3072 | True |
| LRP-Epsilon | Random | 0.7256 | 0.001 | 0.6136 | 0.8375 | True |
| Input * Gradient | LRP-Z | 0.0076 | 0.9 | -0.1044 | 0.1196 | False |
| Input * Gradient | LRP-PresetAFlat | 0.012 | 0.9 | -0.1 | 0.1239 | False |
| Input * Gradient | LRP-AlphaBeta 21 | 0.0258 | 0.9 | -0.0861 | 0.1378 | False |
| Input * Gradient | SmoothGrad | 0.0365 | 0.9 | -0.0755 | 0.1484 | False |
| Input * Gradient | LRP-AlphaBeta 32 | 0.0459 | 0.9 | -0.0661 | 0.1579 | False |
| Input * Gradient | LRP-AlphaBeta 10 | 0.0731 | 0.6431 | -0.0388 | 0.1851 | False |
| Input * Gradient | Gradient | 0.1199 | 0.0222 | 0.0079 | 0.2319 | True |
| Input * Gradient | Deconvnet | 0.1914 | 0.001 | 0.0795 | 0.3034 | True |
| Input * Gradient | Expert | 0.193 | 0.001 | 0.081 | 0.3049 | True |
| Input * Gradient | Random | 0.7232 | 0.001 | 0.6113 | 0.8352 | True |

Table 12 – *Continued from previous page*
| group1 | group2 | meandiff | p-adj | lower | upper | reject |
| --- | --- | --- | --- | --- | --- | --- |
| LRP-Z | LRP-PresetAFlat | 0.0044 | 0.9 | -0.1076 | 0.1163 | False |
| LRP-Z | LRP-AlphaBeta 21 | 0.0182 | 0.9 | -0.0937 | 0.1302 | False |
| LRP-Z | SmoothGrad | 0.0288 | 0.9 | -0.0831 | 0.1408 | False |
| LRP-Z | LRP-AlphaBeta 32 | 0.0383 | 0.9 | -0.0737 | 0.1502 | False |
| LRP-Z | LRP-AlphaBeta 10 | 0.0655 | 0.7938 | -0.0464 | 0.1775 | False |
| LRP-Z | Gradient | 0.1123 | 0.0485 | 0.0003 | 0.2242 | True |
| LRP-Z | Deconvnet | 0.1838 | 0.001 | 0.0719 | 0.2958 | True |
| LRP-Z | Expert | 0.1853 | 0.001 | 0.0734 | 0.2973 | True |
| LRP-Z | Random | 0.7156 | 0.001 | 0.6037 | 0.8276 | True |
| LRP-PresetAFlat | LRP-AlphaBeta 21 | 0.0139 | 0.9 | -0.0981 | 0.1258 | False |
| LRP-PresetAFlat | SmoothGrad | 0.0245 | 0.9 | -0.0875 | 0.1364 | False |
| LRP-PresetAFlat | LRP-AlphaBeta 32 | 0.0339 | 0.9 | -0.078 | 0.1459 | False |
| LRP-PresetAFlat | LRP-AlphaBeta 10 | 0.0612 | 0.8802 | -0.0508 | 0.1731 | False |
| LRP-PresetAFlat | Gradient | 0.1079 | 0.0737 | -0.004 | 0.2199 | False |
| LRP-PresetAFlat | Deconvnet | 0.1795 | 0.001 | 0.0675 | 0.2914 | True |
| LRP-PresetAFlat | Expert | 0.181 | 0.001 | 0.069 | 0.2929 | True |
| LRP-PresetAFlat | Random | 0.7113 | 0.001 | 0.5993 | 0.8232 | True |
| LRP-AlphaBeta 21 | SmoothGrad | 0.0106 | 0.9 | -0.1013 | 0.1226 | False |
| LRP-AlphaBeta 21 | LRP-AlphaBeta 32 | 0.0201 | 0.9 | -0.0919 | 0.132 | False |
| LRP-AlphaBeta 21 | LRP-AlphaBeta 10 | 0.0473 | 0.9 | -0.0647 | 0.1593 | False |
| LRP-AlphaBeta 21 | Gradient | 0.0941 | 0.2249 | -0.0179 | 0.206 | False |
| LRP-AlphaBeta 21 | Deconvnet | 0.1656 | 0.001 | 0.0537 | 0.2776 | True |
| LRP-AlphaBeta 21 | Expert | 0.1671 | 0.001 | 0.0552 | 0.2791 | True |
| LRP-AlphaBeta 21 | Random | 0.6974 | 0.001 | 0.5855 | 0.8094 | True |
| SmoothGrad | LRP-AlphaBeta 32 | 0.0094 | 0.9 | -0.1025 | 0.1214 | False |
| SmoothGrad | LRP-AlphaBeta 10 | 0.0367 | 0.9 | -0.0753 | 0.1486 | False |
| SmoothGrad | Gradient | 0.0834 | 0.4347 | -0.0285 | 0.1954 | False |
| SmoothGrad | Deconvnet | 0.155 | 0.001 | 0.043 | 0.267 | True |
| SmoothGrad | Expert | 0.1565 | 0.001 | 0.0445 | 0.2685 | True |
| SmoothGrad | Random | 0.6868 | 0.001 | 0.5748 | 0.7988 | True |
| LRP-AlphaBeta 32 | LRP-AlphaBeta 10 | 0.0272 | 0.9 | -0.0847 | 0.1392 | False |
| LRP-AlphaBeta 32 | Gradient | 0.074 | 0.6261 | -0.038 | 0.186 | False |
| LRP-AlphaBeta 32 | Deconvnet | 0.1456 | 0.001 | 0.0336 | 0.2575 | True |
| LRP-AlphaBeta 32 | Expert | 0.1471 | 0.001 | 0.0351 | 0.259 | True |
| LRP-AlphaBeta 32 | Random | 0.6774 | 0.001 | 0.5654 | 0.7893 | True |
| LRP-AlphaBeta 10 | Gradient | 0.0468 | 0.9 | -0.0652 | 0.1587 | False |
| LRP-AlphaBeta 10 | Deconvnet | 0.1183 | 0.0263 | 0.0063 | 0.2303 | True |
| LRP-AlphaBeta 10 | Expert | 0.1198 | 0.0224 | 0.0079 | 0.2318 | True |
| LRP-AlphaBeta 10 | Random | 0.6501 | 0.001 | 0.5381 | 0.7621 | True |
| Gradient | Deconvnet | 0.0716 | 0.6746 | -0.0404 | 0.1835 | False |
| Gradient | Expert | 0.0731 | 0.6447 | -0.0389 | 0.185 | False |
| Gradient | Random | 0.6034 | 0.001 | 0.4914 | 0.7153 | True |
| Deconvnet | Expert | 0.0015 | 0.9 | -0.1105 | 0.1135 | False |
| Deconvnet | Random | 0.5318 | 0.001 | 0.4198 | 0.6438 | True |
| Expert | Random | 0.5303 | 0.001 | 0.4183 | 0.6423 | True |

**Table 13:**
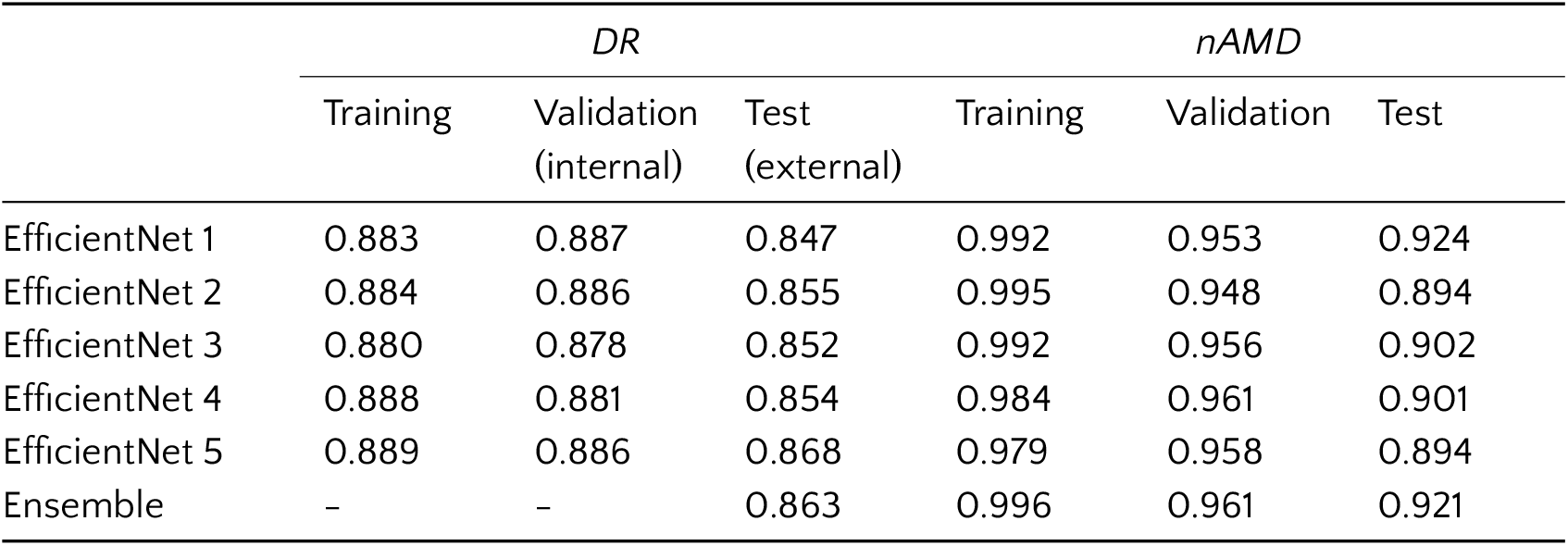
Disease detection accuracy for EfficientNets and their ensembles.

We also performed perturbation analyses [11, 76, 44] and compared the perturbation trajectory of saliency maps to those of clinicians in order to obtain an alternative perspective on the clinical relevance of saliency maps. Our perturbation scheme involved a two-dimensional grid specified over a given image and we regarded each cell as a patch to be perturbed (Fig. 6). Then, given a saliency map, we ranked the patches based on the total patch saliency, replaced the top-ranked patches with uniform random noise as per [76] and measured the drop in the *ensemble* output for the class of interest, `diseased`. We followed the ranking and repeated the measurement until there was no more patch to replace. In addition, we used random maps to facilitate random perturbation as our baseline. As expected, a saliency-based ranking led to faster decline than a random selection of patches, since the saliency map indicated the informative regions in an image more accurately than chance. Analogously, we used the rate of drop as a performance metric for saliency maps. However, when the total perturbation grew and disease evidence was lost, all methods converged to random (Fig. 8a and Fig. 8b). After all, we treated also expert annotations as saliency maps within this perturbation-based framework. As clinical annotations were represented in terms of 1s (region of interest) or 0s (background), the patch ranking in this case was achieved according to the sum of 1s in patches. If the patch size was smaller than the annotated area and there were multiple patches full of 1s, the ordering of these patches did not really matter on average. Once the patches with equal number of 1s were visited, the remaining patches were selected based on their sums. Thanks to the numerical representation of expert annotations, our procedures for perturbation analysis were readily applicable to them. This allowed us to validate saliency maps against clinicians by monitoring DNNs’ sensitivity to the removal of salient information determined by explanation methods as well as the clinicians themselves.

**Figure 6:**
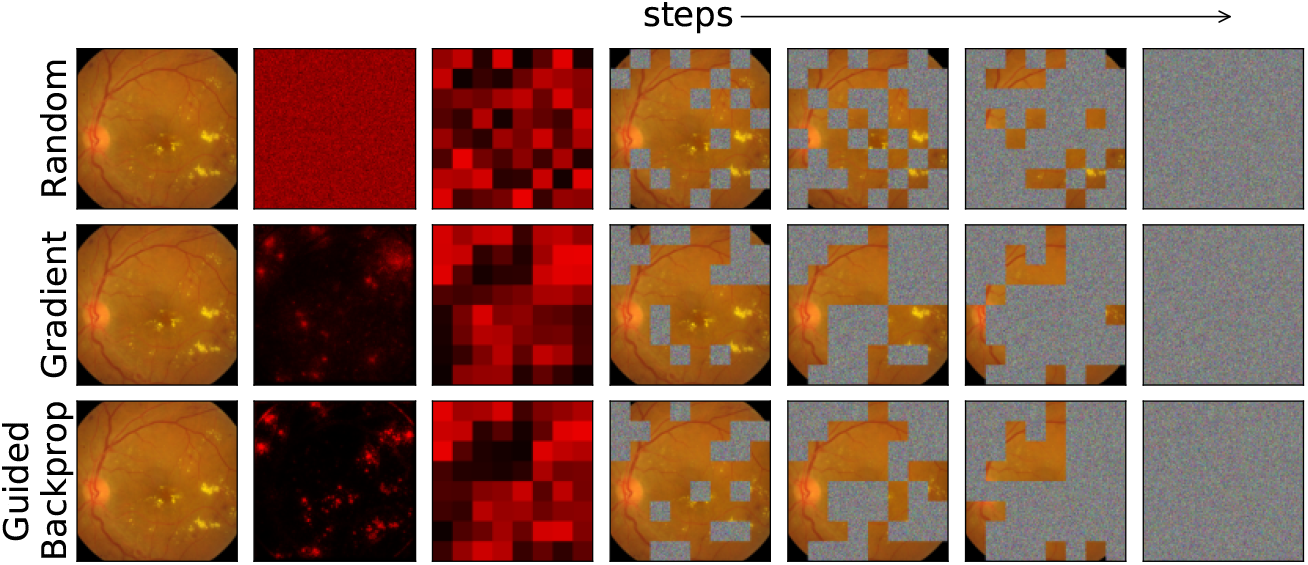
Illustration of perturbation analysis. Given a fundus image with DR (the first column) and three saliency maps (the second column) for it, 64 *×* 64 patches lead to 8 *×* 8 grids (the third column) with different rankings of patches. If 16 patches are perturbed per step, the image is fully perturbed in 4 steps.

**Figure 7:**
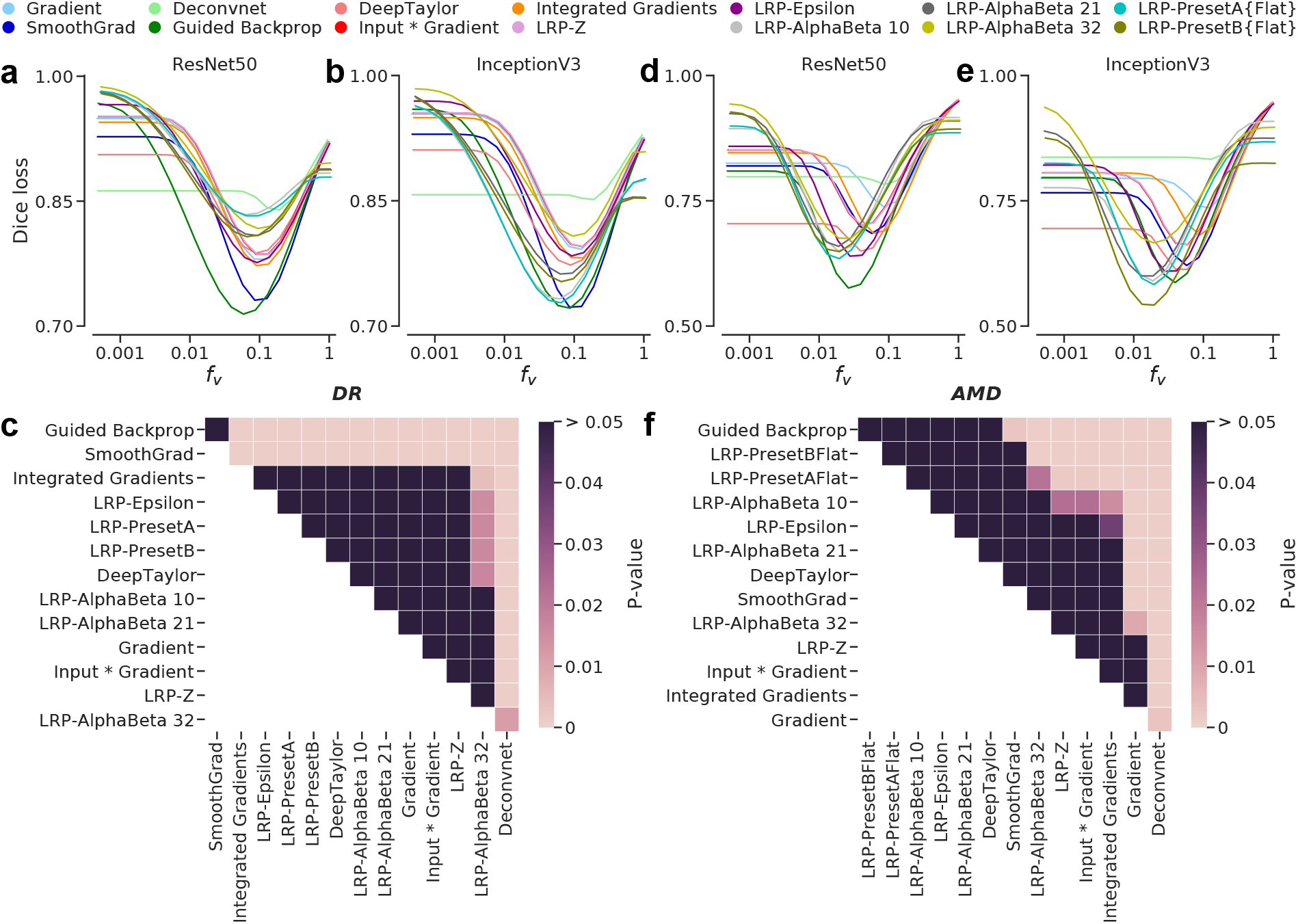
Comparison of ensemble-based saliency maps with expert annotations. Curves indicate the mean Dice loss between saliency maps and expert annotations. Multiple comparisons of attribution methods based on the minimum mean Dice loss for the overall DR and nAMD scenarios are given in grids with cell colors indicating significance. Rows and columns are ordered in an ascending fashion w.r.t. the minimum mean Dice loss achieved by methods. **(a-c)** Results for the DR detection task with expert annotations *excluding the optic disc*. See Fig. 12 in Appendix A.3 for curves w.r.t. annotations of individual lesions, full annotation including the optic disc as well as the unannotated regions. **(d-f)** Results for the nAMD activity detection task with complete expert annotations. See Fig. 13 in Appendix A.3 for curves w.r.t. annotations of intraretinal or subretinal fluid as well as the unannotated regions.

**Figure 8:**
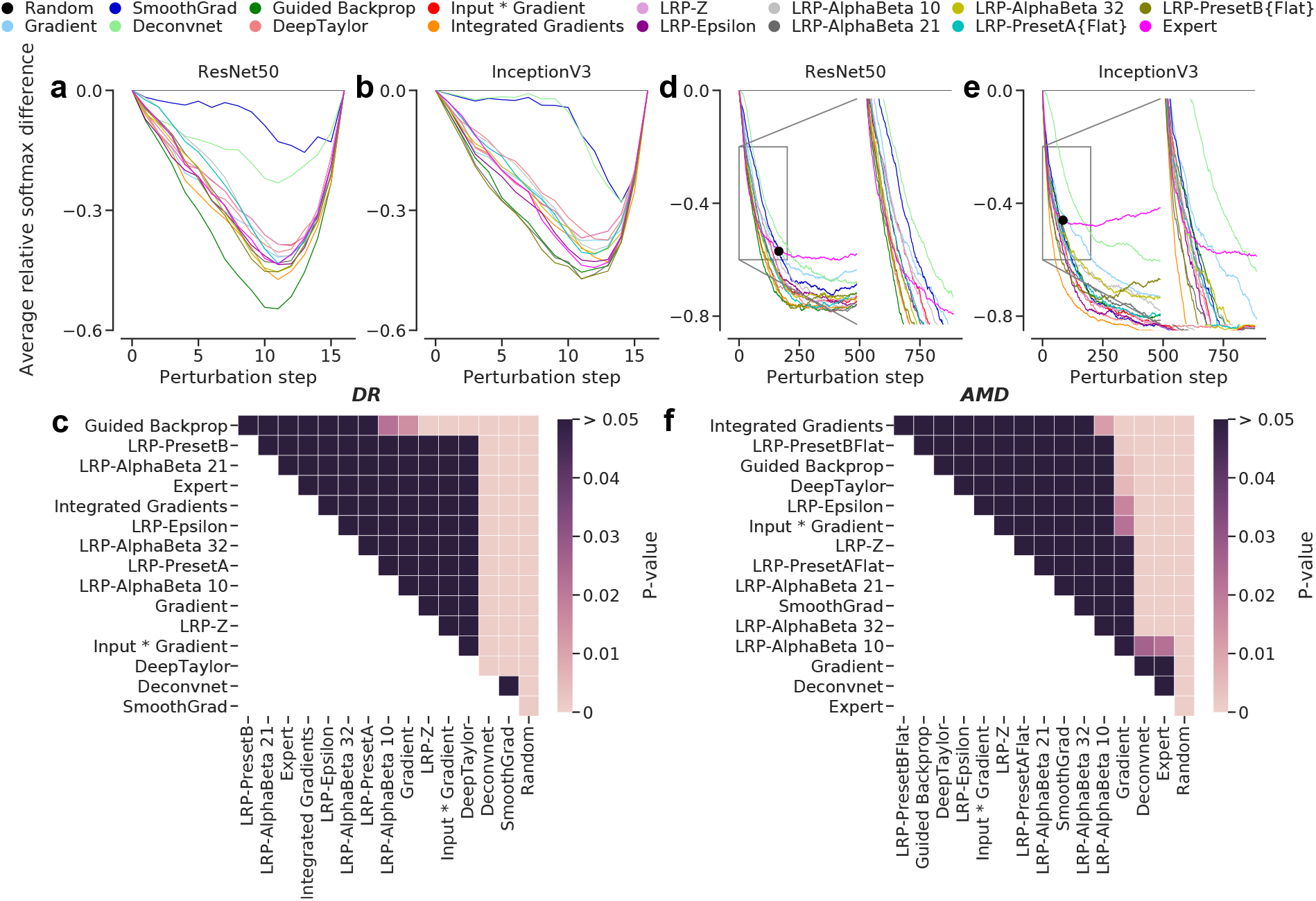
Perturbation analyses including the expert annotations as saliency maps. Curves were obtained by measuring the average differences from the *random* baseline. Thus, the baseline is shown as a flat line and all other methods converge to it, as the total perturbation grows and evidence is lost. Multiple comparisons of attribution methods based on the relative differences at steps 10 and 200, for the overall DR and nAMD scenarios, respectively, are given in grids with cell colors indicating significance. Rows and columns are ordered in an ascending fashion w.r.t. the relative differences achieved by methods. **(a-c)** Results for the DR detection task w.r.t. expert annotations *excluding the optic disc*. **(d-f)** Results for the nAMD activity detection task with complete expert annotations. The insets in **(d)** and **(e)** focus on the steps between 0 and 200 inclusively. Black dots indicate the points of divergence between the expert and methods.

For fundus images, which were accompanied with widespread annotations, we used the settings described in Fig. 6 but we perturbed 4 patches per step. Thus, a fundus image was fully perturbed in 16 steps (Fig. 8a and Fig. 8b). Considering the local annotations of retinal fluid on B-scans, we increased the granularity of perturbations in order to precisely monitor the changes in the DNN outputs for nAMD activity. We used patches of 4 *×* 4 on a grid of 110 *×* 128 and perturbed 4 patches per step. To sidestep the formidable computation required to run the full-fledged analyses for this task, we stopped early after the 880^th^ step out of 3520 (Fig. 8d and Fig. 8e). After all, we plotted the average relative differences in the ensemble outputs for being diseased against the steps (Fig. 8), by subtracting the drop observed via a random perturbation from those of ranked perturbations. As a performance metric, we used the values induced by attribution methods at steps 10 and 200 for the DR and nAMD scenarios, respectively.

## Results

We developed DNNs to detect DR and active nAMD from retinal fundus images (Fig. 1a) and slices of OCT volume scans (Fig. 1c), respectively. For each disease, we mainly used two well-known network architectures, ResNet50 [38] and InceptionV3 [86], as well as a newer one: EfficientNet [87], in particular, Efficient-Net (B5) – the largest variant that we could fit into our computational pipeline. Then, we constructed two Deep Ensembles [47, 29] for each diagnostic task, which each consisted of five DNNs from a given architecture, trained with different random initializations and data augmentation. Thus, we used 30 DNNs in this study. While individual DNNs were accurate for their respective tasks, their ensembles further improved upon single network performance in both disease scenarios and across network architectures (Table 3 and Fig. 2 as well as Table 13 and Fig. 14 in Appendix A.5). We also assessed the calibration of our ensembles via reliability diagrams and the Adaptive Expected Calibration Error (AECE) [22] and found them to be well calibrated (Fig. 2 and Fig. 14). For the sake of brevity and clarity, we focus on the ResNet50 and InceptionV3 architectures and present the EfficientNet results mostly in Appendix A.5.

Interestingly, the diversity of DNNs in decision-making showed clearly in saliency maps. For example, the first two DR detection networks paid more attention to the hemorrhages, microaneurysm (indicated by a dotted arrow) and soft exudates (bottom right, Fig. 3a), while the soft exudate was completely unattended by the last three DNN instances. The fifth one also ignored hemorrhages and detected only microaneurysm in this area. In addition to the annotated lesions, the DNNs also detected two hemorrhages (indicated by solid arrows) at the bottom left (for more examples, see the first two rows of Fig. 10 in Appendix A.2). Similarly, the nAMD activity detection networks used the presence of intraretinal or subretinal fluid as revealed by saliency maps (Fig. 3b). However, the first DNN did not pay much attention to the subretinal fluid, while the fifth one highlighted it along with additional intraretinal cues. Despite the differences, DNNs also agreed on the saliency of the top end of the large intraretinal lesion. After all, the ensembles of DNNs led to well-informed and comprehensive saliency maps, thanks to the aggregation of different views from individual DNNs (Fig. 3). However, even the ensemble-based saliency maps were not immediately amenable to human interpretation, as they were extremely sparse (Fig. 5, top rows in (a) and (b)). We used a custom-developed post-processing method (see Methods) to improve the visualization of salient regions (Fig. 5). It also normalized the saliency scores that varied wildly due to the differences between attribution methods and network architectures.

We used such enhanced ensemble-based saliency maps to systematically evaluate the clinical relevance of DNNs with a focus on explainability. We first compared the saliency maps with expert annotations (Fig. 7), which were presented as segmentation maps (Fig. 1b and Fig. 1d), and assessed their (dis)similarities directly via Dice loss [58]. To exclude potentially misleading saliency maps due to misclassification from the analysis, we considered only the images that were correctly classified by all members of respective ensembles. Interestingly, all annotated fundus images from the IDRiD collection were correctly classified by all DR detection networks. This is likely due to the severity and spread of lesions in these images. For nAMD activity detection, DNNs with ResNet50 and InceptionV3 architectures classified 62 and 55 B-scans (out of 71 with expert annotation) correctly, respectively. In order to obtain balanced groups for our analysis, we considered the intersection of these two sets containing 52 B-scans.

We used the optimally post-processed saliency maps for each combination of disease scenario, DNN architecture and attribution method (see Methods) and asked whether the match of the saliency maps to the clinical annotation was significantly influenced by DNN architecture or the attribution method (2-way repeated measures ANOVA, see Appendix A.4 for details). In the DR detection task, DNN architecture (F(1,80) = 41.340, p = 8.6 × 10^−9^) and attribution method (F(13,1040) = 43.764, p = 3.0 × 10^−89^) as well as their interaction (F(13,1040) = 106.684, p = 6.2 × 10^−181^) had a significant influence. We obtained similar results for the nAMD activity detection task (F(1,51) = 65.573, p = 1.0 × 10^−10^ and F(13,663) = 29.354, p = 3.8 × 10^−57^ for the main effects and F(13,663) = 44.823, p = 6.6 × 10^−82^ for their interaction). Using posthoc testing, we found significant pairwise differences between the mean Dice loss of different attribution methods: For DR detection (Fig. 7a and b), Guided Backprop and SmoothGrad were competitive with each other and significantly outperformed all other methods (Fig. 7c). Guided Backprop also performed well in the nAMD activity detection task (Fig. 7d and e). It outperformed seven methods including SmoothGrad (Fig. 7f). However, five LRP configurations along with Deep Taylor were as good as Guided Backprop on average in this task. After all, DeConvNet yielded the worst saliency maps in terms of the match to clinical annotations.

We next studied which kind of lesions were most strongly highlighted in saliency maps, indicating that they play a key role in the diagnostic decisions of DNNs. For DR, we found that DNNs relied more on small lesions, such as microaneurysms (green) and hard exudates (dark blue), but they typically captured them incompletely (Fig. 10 in Appendix A.2). In contrast, large instances of soft exudates (cyan) and hemor-rhages (magenta) were less taken into account by the DNNs. Even when such large lesions were attended by DNNs, they were only partially covered in saliency maps. As a result, the Dice loss for individual lesion types was larger on average for soft exudates than hard exudates, for example, but that strongly differed between methods (Fig. 12e-h in Appendix A.3). Likewise, substantially large hemorrhages were almost completely ignored by DNNs (Fig. 10, 4th row). Also, different saliency methods highlighted different lesions or anatomical structures in the retina, even for the same network architecture (Fig. 10). For instance, Guided Backprop almost always pointed at DR lesions, whereas SmoothGrad often focused on vessels (in and out of the optic disc) and captured fewer lesions. While Guided Backprop’s top preferences were microaneurysms and hemorrhages (Fig. 12a-d), hard and soft exudates as well as the optic disc were typical formations highlighted by SmoothGrad (Fig. 12e-j). Integrated Gradients also behaved similar to Guided Backprop but it performed worse than the two overall. Finally, we observed that our post-processing method emphasized not only the lesions themselves but also their surroundings. In particular, tiny lesions such as microaneurysms and hard exudates were subject to overgrowing in saliency maps, since we tuned *f*_*v*_ with respect to Dice loss on the complete set of annotations, including those for large lesions. As a result, the average Dice loss values for microaneurysms and hard exudates were increased on top of these lesions being captured incompletely in the first place (Fig. 12a,b,e and f in Appendix A.3). This combined with the errors made on different parts reduced the overall gap between Guided Backprop and Smooth-Grad (Fig. 7a-c), even though their saliency maps looked quite different. On the bright side, our method was effective at detecting tiny relevance scores in the vicinity of DR lesions and bringing them up to human attention. In the nAMD activity detection task, small retinal fluid were the *go-to* pathology for DNNs (Fig. 11 in Appendix A.2). However, the large ones were not ignored, either. DNNs typically responded to the boundaries of large retinal fluid and saliency maps showed a cavity in the interior (Fig. 11, last three rows). Thus, the Dice loss for intraretinal fluid was larger than for subretinal fluid on average (Fig. 13 in Appendix A.3), since the former was usually larger in size than the latter. Interestingly, saliency methods were more consistent about their preferences for salient regions in this case. We attribute this to the small variety of pathologies. However, in addition to retinal fluid, DNNs used features from the fovea to discern nAMD activity (Fig. 11), even though it was not annotated by experts as key for the task. On the other hand, the effect of our post-processing was again apparent in saliency maps (Fig. 11). The retinal fluid and their surroundings were highlighted together and the Dice loss for small subretinal fluid was high on average (Fig. 13).

Next, we used perturbation analysis to validate the optimal saliency maps with respect to expert annotations. To this end, we used the expert annotations of clinically relevant lesions also as saliency maps. We performed 2-way repeated measures ANOVA based on the average differences between the ensemble outputs induced by ranked and random perturbations using the aforementioned design. In the DR detection task, we found that DNN architecture did not significantly influence our measure (F(1,80) = 1.901, p = x 10^−1^), whereas the choice of attribution method had a significant effect (F(15,1200) = 113.691, p = 7.8 × 10^−218^) as had interaction of these two factors (F(15,1200) = 5.466, p = 7.8 × 10^−11^). The effects followed a similar trend in the nAMD activity detection task (main effects: F(1,51) = 0.189, p = 6.7 × 10^−1^; F(15,765) = 116.869, p = 4.2 × 10^−186^); interaction: F(15,765) = 6.004, p = 5.8 × 10^−12^). Using post-hoc testing, we again found significant pairwise differences between the means of attribution methods. In the DR detection task (Fig. 8a and Fig. 8b), Guided Backprop was the best method on average, competitive with seven methods, including the expert annotation, and significantly outperforming eight methods (Fig. 8c). Also, the expert annotation performed not significantly different than a number of saliency methods and better than SmoothGrad and DeConvNet on average. In the nAMD activity detection task (Fig. 8d and Fig. 8e), saliency methods and expert annotation closely followed in the early stages of perturbations. However, the expert curves quickly stabilized into almost flat lines. The flat lines indicated that the perturbation order essentially followed random selection of patches once the most important pathologies annotated by clinicians were removed. Perturbations with respect to saliency maps led to further reduction beyond the expert curves, indicating the use of additional features by DNNs. After all, Integrated Gradients outperformed five methods, one of which was the expert annotation (Fig. 8f). Guided Backprop, Deep Taylor, Input *×* Gradient, SmoothGrad and six LRP configurations were as good as Integrated Gradients on average. Surprisingly, DeConvNet achieved a better performance in comparison with the earlier scenarios.

Our two analyses — direct comparisons of lesions using Dice loss and perturbation analysis — provided complementary information about the factors influencing the quality of saliency maps: The first analysis indicated that the DNN architecture can be a role for explainability, interacting with the attribution method. Across tasks and network architectures, Guided Backprop emerged as the most useful method for generating clinically relevant saliency maps (Fig. 7). Also, the methods, e.g., Guided Backprop and SmoothGrad, differed in their preferences for salient lesions and anatomical structures in the retina, even for a given architecture. For the perturbation analysis, we did not find an effect of DNN architecture and we observed similarities between the perturbation trajectories of many saliency methods and expert annotations (Fig. 8). The use of large patches combined with the spread and severity of DR lesions probably suppressed the differences between DNNs and clinicians in DR detection (Fig. 8a-b). But, in the nAMD scenario, the trajectories of saliency methods and expert annotation diverged after an initial period of collective descent (Fig. 8d-e). Interestingly, the curves based on the saliency methods continued to descent past the expert curves, suggesting that a few key instances of retinal fluid were mostly enough for a clinician to make a diagnosis, while DNNs also used fovea characteristics for detecting nAMD activity.

In follow-up to the above analyses, we examined the B-scans that led to disagreement among clinicians or DNNs over nAMD activity. First, we looked at two images with different diagnoses made by our clinical experts (Fig. 9a-b). These were initially graded as *inactive*; however, tiny instances of subretinal fluid were detected by another expert during pixel-level annotation. As DNNs were trained with the image-level grades, they mimicked the main grader’s diagnoses. But, saliency maps indicated that DNNs also found some evidence in and around the annotated regions, thanks to our processing method’s ability to highlight tiny regions of relevance. The evidence accumulated by DNNs, however, was not enough to make them predict active nAMD in both images. In the next three cases (Fig. 9c-e), our clinical experts agreed on the disease activity, whereas DNNs demonstrated disagreement among themselves. On the bright side, ensemble decisions were correct and associated with well-calibrated probabilities (Fig. 2) to indicate the predictive uncertainty for these examples. The uncertainty information can be used to judge these automated diagnoses [13, 28, 33, 34] or to refer the cases to other physicians for further inspection [49, 9, 10]. Surprisingly, DNNs also occasionally disagreed on consecutive slices (Fig. 9f-g, 17th and 18th slices) of a given volume scan. High uncertainty (low confidence) of the ensemble decisions and the disagreement of saliency maps were indicating the dangers of automated decisions here. Interestingly, DNNs made errors on obvious cases, as well (Fig. 9h). Their previously discussed decision mechanisms, such as focusing on lesion boundaries or relatively small structures, showed again in this particular example but only 1 DNN made a wrong prediction with high uncertainty (probability of 0.45). Overall, saliency maps also showed that DNNs with matching decisions were typically more compatible with each other, compared to those with disagreement (Fig. 9, last column). This indicated that saliency maps can be used to detect and resolve disagreements among DNNs as well as clinicians.

**Figure 9:**
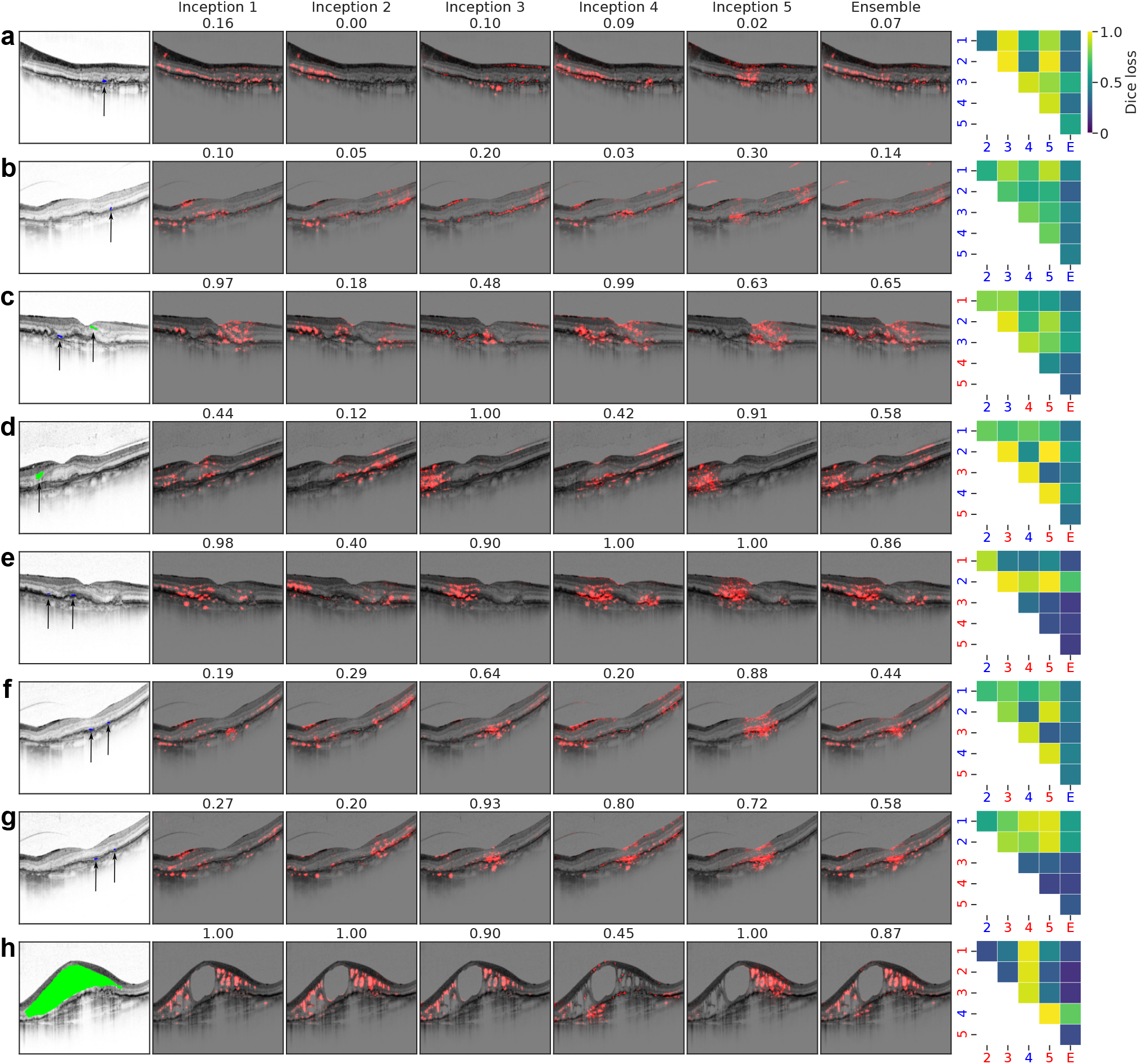
Disagreement among clinicians as well as DNNs over nAMD activity. Exemplary B-scans with active nAMD and expert annotations of retinal fluid are given in the first column. Arrows indicate retinal fluid. Coloring of retinal fluid is same as in Fig. 1d. Best viewed in color and when zoomed in. These images led to disagreement among clinicians **(a-b)** or ensemble members **(c-h)**. Saliency maps in columns 2-7 were obtained via LRP-PresetBFlat from the InceptionV3 models or their ensemble. Our processing method with the corresponding optimal *f*_*ν*_ was also applied. Predictive probabilities for nAMD activity are given on top of saliency maps. The last column shows pairwise Dice loss among saliency maps. Diagnostics decisions based on predictive probabilities are also indicated via red (active) or blue (inactive).

**Figure 10:**
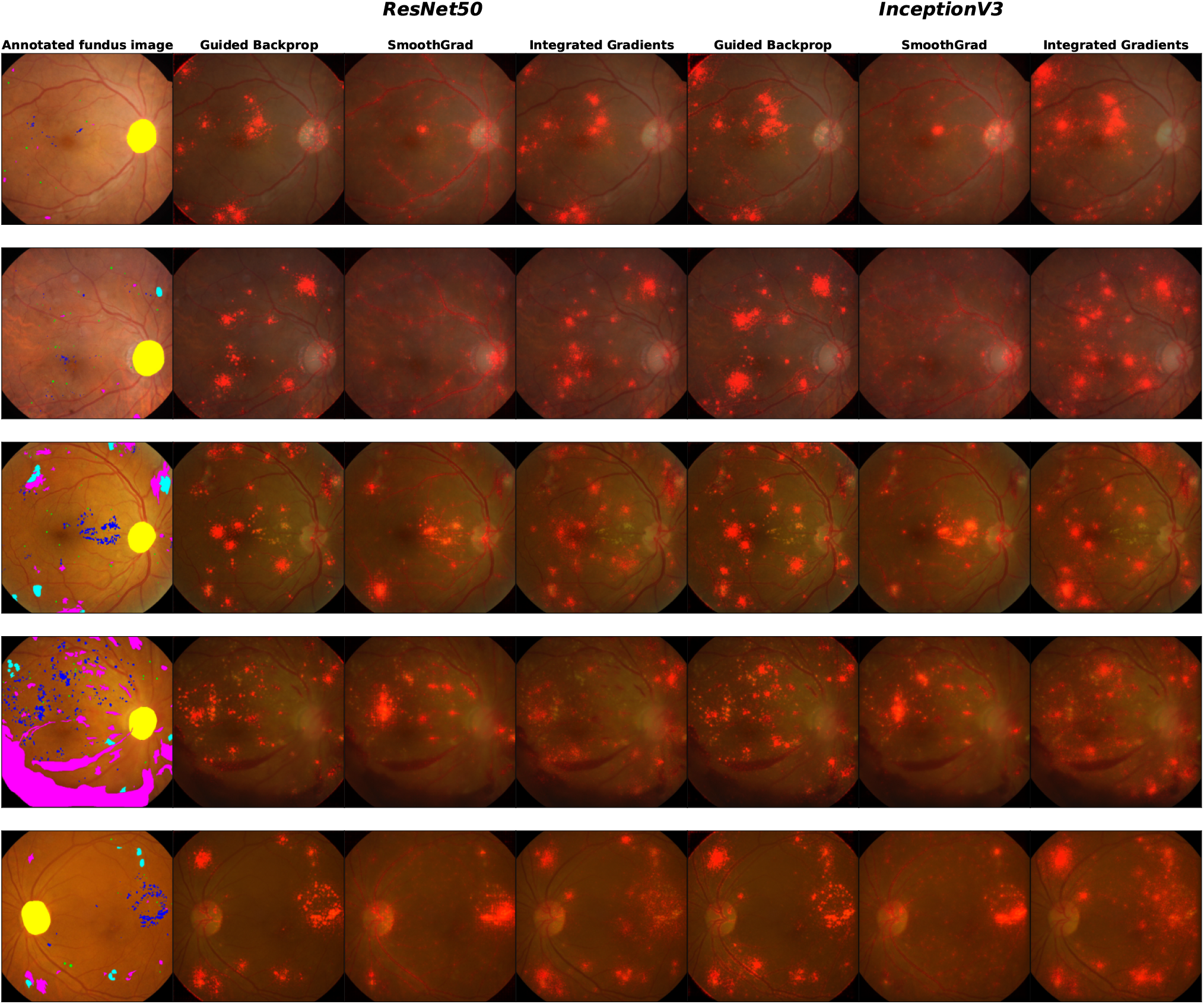
Exemplary saliency maps obtained via our processing method and the best *f*_*ν*_ values for the top three attribution methods for the DR detection task. The leftmost column shows fundus images with expert annotations for the pathologies of DR. Coloring of annotations is the same as in Fig. 1. The remaining columns show the ensemble-based saliency maps in two DNN groups.

**Figure 11:**
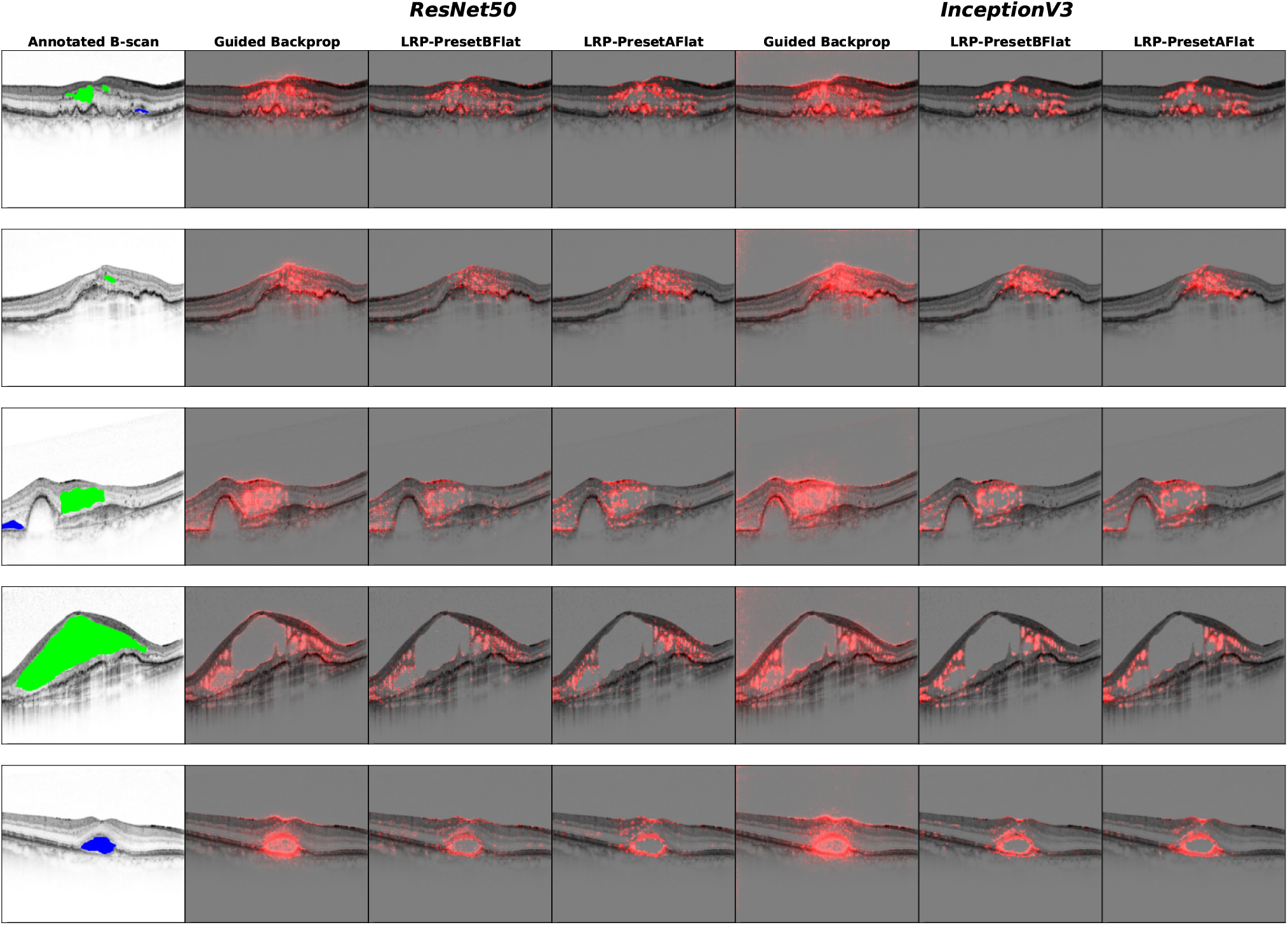
Exemplary saliency maps obtained via our processing method and the best *f*_*ν*_ values for the top three attribution methods for the AMD activity detection task. The leftmost column shows AMD-active B-scans with expert annotations for retinal fluid. Coloring of annotations is the same as in Fig. 1. The remaining columns show the ensemble-based saliency maps in two DNN groups.

**Figure 12:**
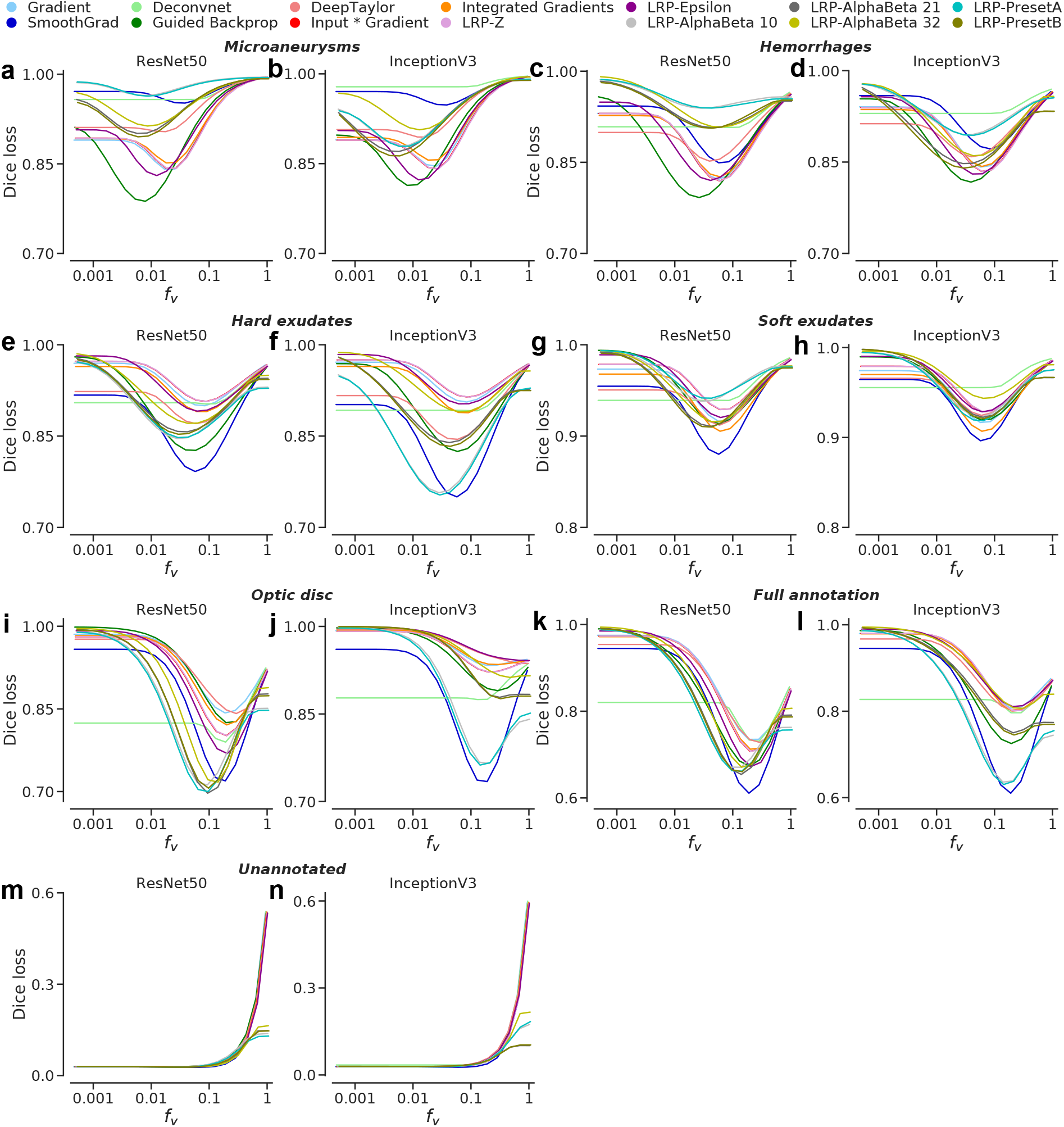
Comparison of ensemble-based saliency maps with expert annotations w.r.t individual lesions, full annotation including the optic as well as the unannotated regions. Curves indicate the mean Dice loss between saliency maps and expert annotations. **(a-b)** Microaneurysms (MA) **(c-d)** Hemorrhages (HE) **(e-f)** Hard exudates (EX) **(g-h)** Soft exudates (SE) **(i-j)** The optic disc (OD) **(k-l)** Full set of annotations including the optic disc **(m-n)** Unannotated regions via the negation of the full set of annotations.

**Figure 13:**
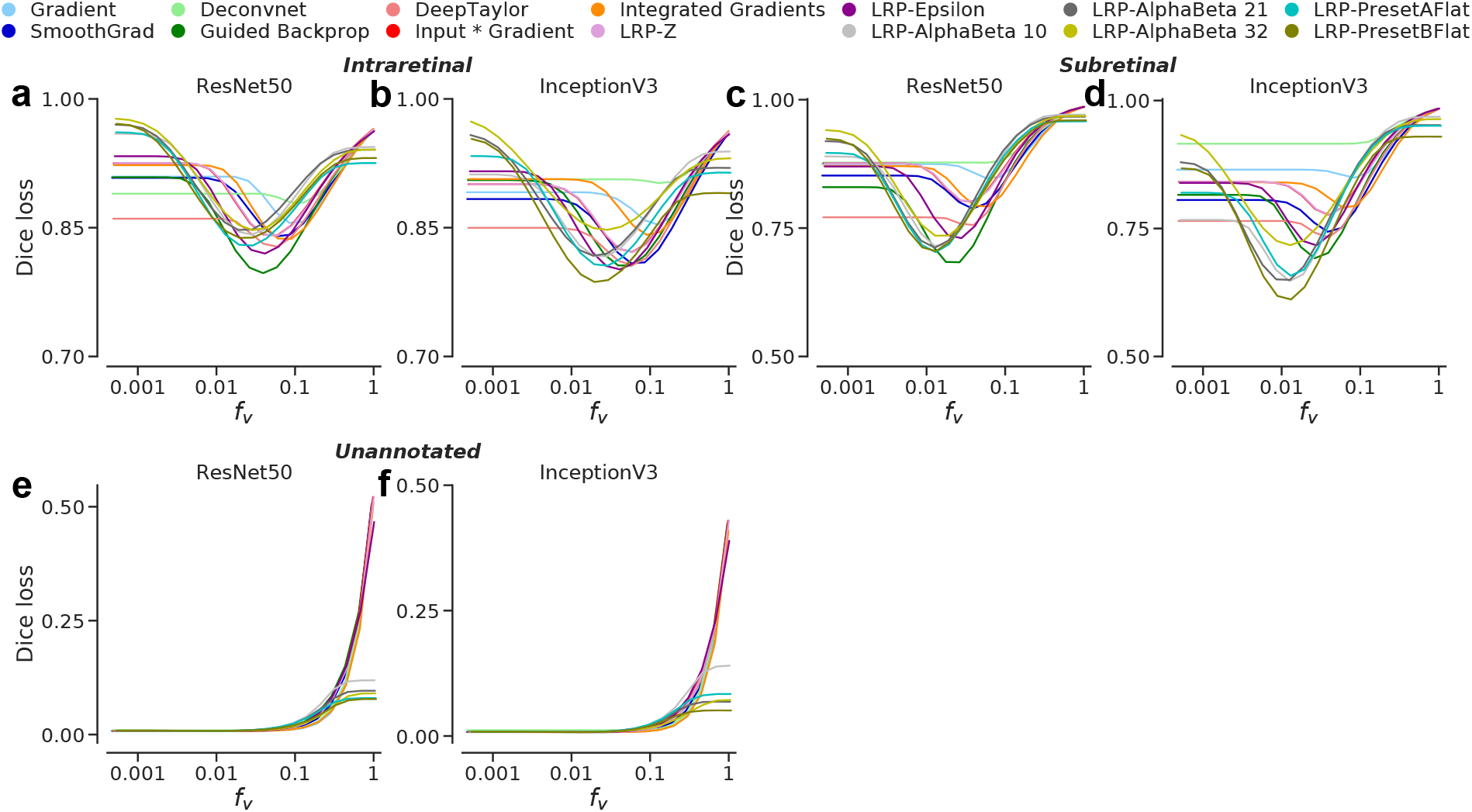
Comparison of ensemble-based saliency maps with expert annotations w.r.t individual lesions as well as the unannotated regions. Curves indicate the mean Dice loss between saliency maps and expert annotations. **(a-b)** Intraretinal fluid **(c-d)** Subretinal fluid **(e-f)** Unannotated regions via the negation of the full set of annotations.

Finally, we revisited our analyses with EfficientNet [87] (Appendix A.5). In terms of disease detection performance, EfficientNet was comparable to ResNet50 and InceptionV3 (Table 13). However, the Efficient-Net ensembles were better calibrated in both disease scenarios (Fig. 14). On the other hand, the Efficient-Net architecture led to compatibility issues with several saliency methods. For instance, EfficientNets use *swish* activation [72, 23], whereas GuidedBackprop is essentially designed for ReLU networks (see Methods). Similarly, EfficientNets use the depth-wise convolutions but these are not supported by the Deep Taylor decomposition implementation in the iNNvestigate toolbox. In addition to these, LRP-Z and LRP-AphaBeta32 rules led to numerical errors. They often generated *NaNs* in raw saliency maps. Therefore, we removed Deep Taylor, LRP-Z and LRP-AlphaBeta32 from the list of saliency methods and performed the same analyses with EfficientNets, but with 50 B-scans (instead of 52) correctly classified by all 30 DNN instances. However, due to the substantially larger memory requirements of EfficientNets, we had to reduce the number of samples (or steps) used for SmoothGrad and Integrated Gradients. We used 16 and 128 samples in the DR and nAMD scenarios, respectively. Despite the relaxation of the ReLU assumption, GuidedBackprop generated saliency maps without numerical errors; however, it lost its clear advantage over other methods in both DR and nAMD scenarios (Fig. 15 and Fig. 16). Interestingly, it was still competitive with other gradient-based methods. But, all LRP configurations were adversely affected by the architectural changes here. They performed worst in both overlap and perturbation analyses.

**Figure 14:**
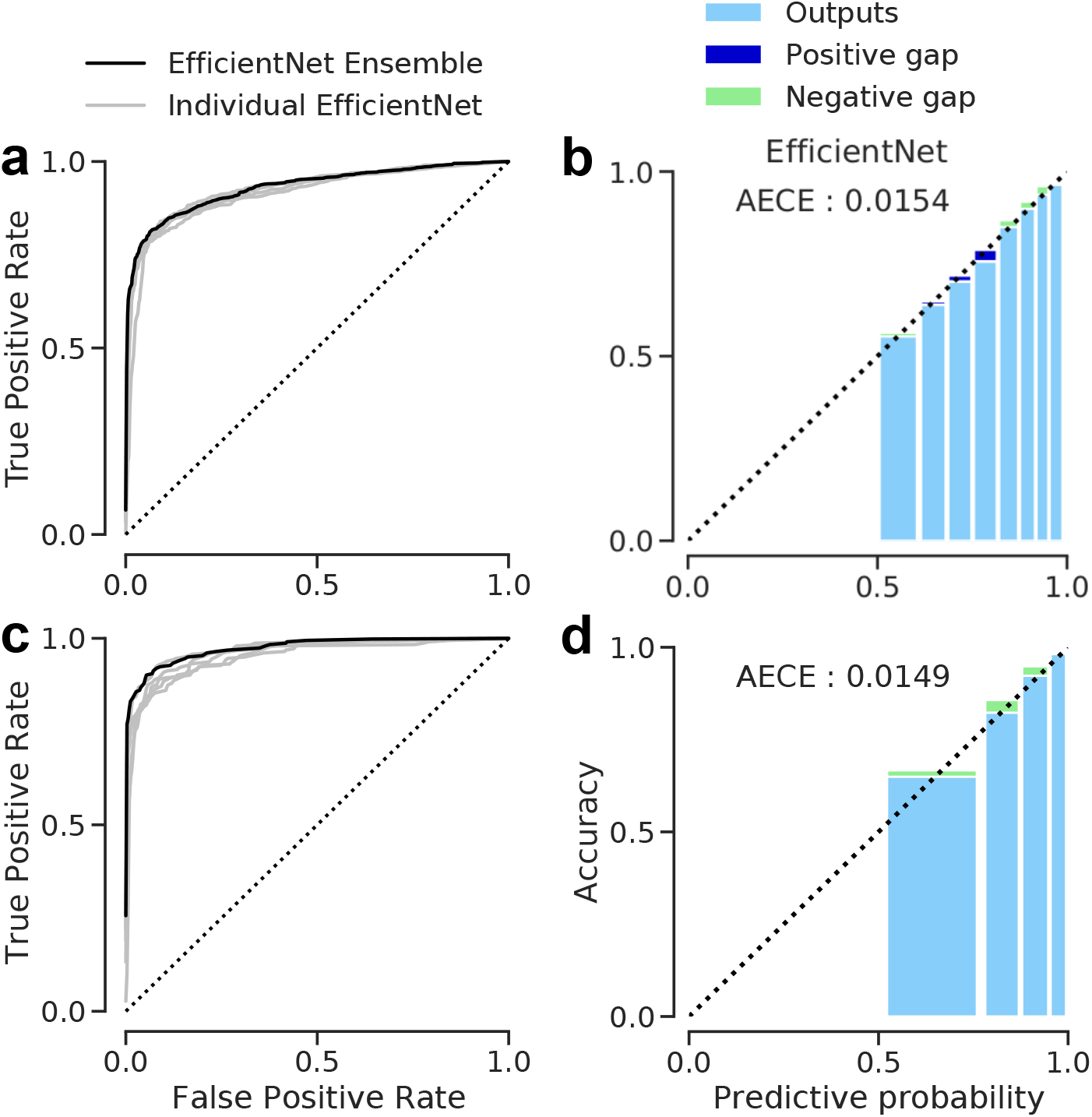
Receiver Operating Characteristics (ROCs) and calibration of the EfficientNet ensembles. **(a-b)**: DR detection. For the sake of clarity, only the performances on external validation set are shown. **(c-d)**: nAMD activity detection. Only the test set performances are shown.

**Figure 15:**
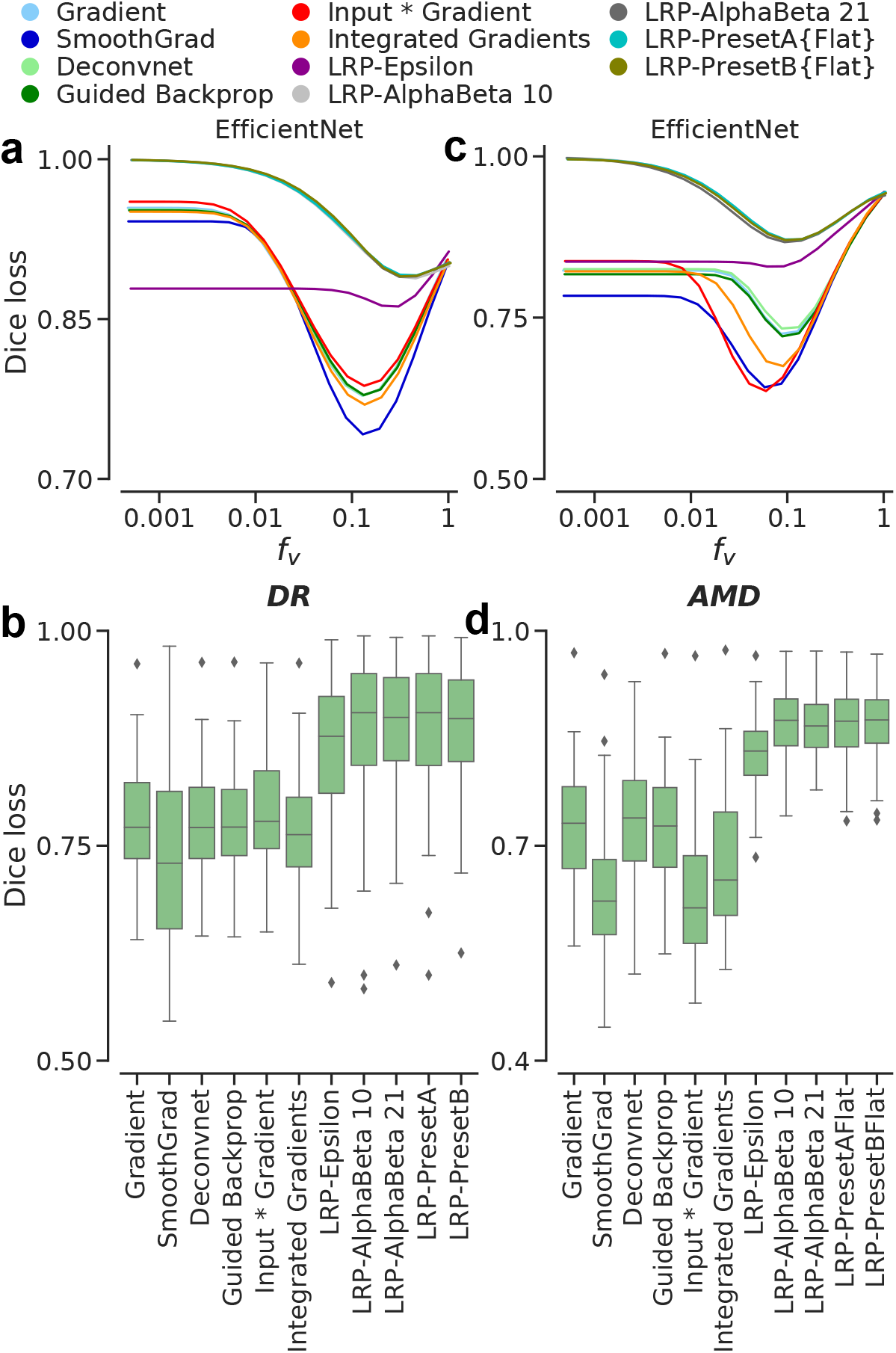
Comparison of ensemble-based saliency maps with expert annotations. Deep Taylor, LRP-Z and LRP-*α*_3_*β*_2_ were removed due to computational issues. Curves indicate the mean Dice loss between saliency maps and expert annotations. Box plots show the distributions of Dice losses for attribution methods. **(a-b)** Results for the DR detection task with expert annotations *excluding the optic disc*. **(c-d)** Results for the nAMD activity detection task with complete expert annotations.

**Figure 16:**
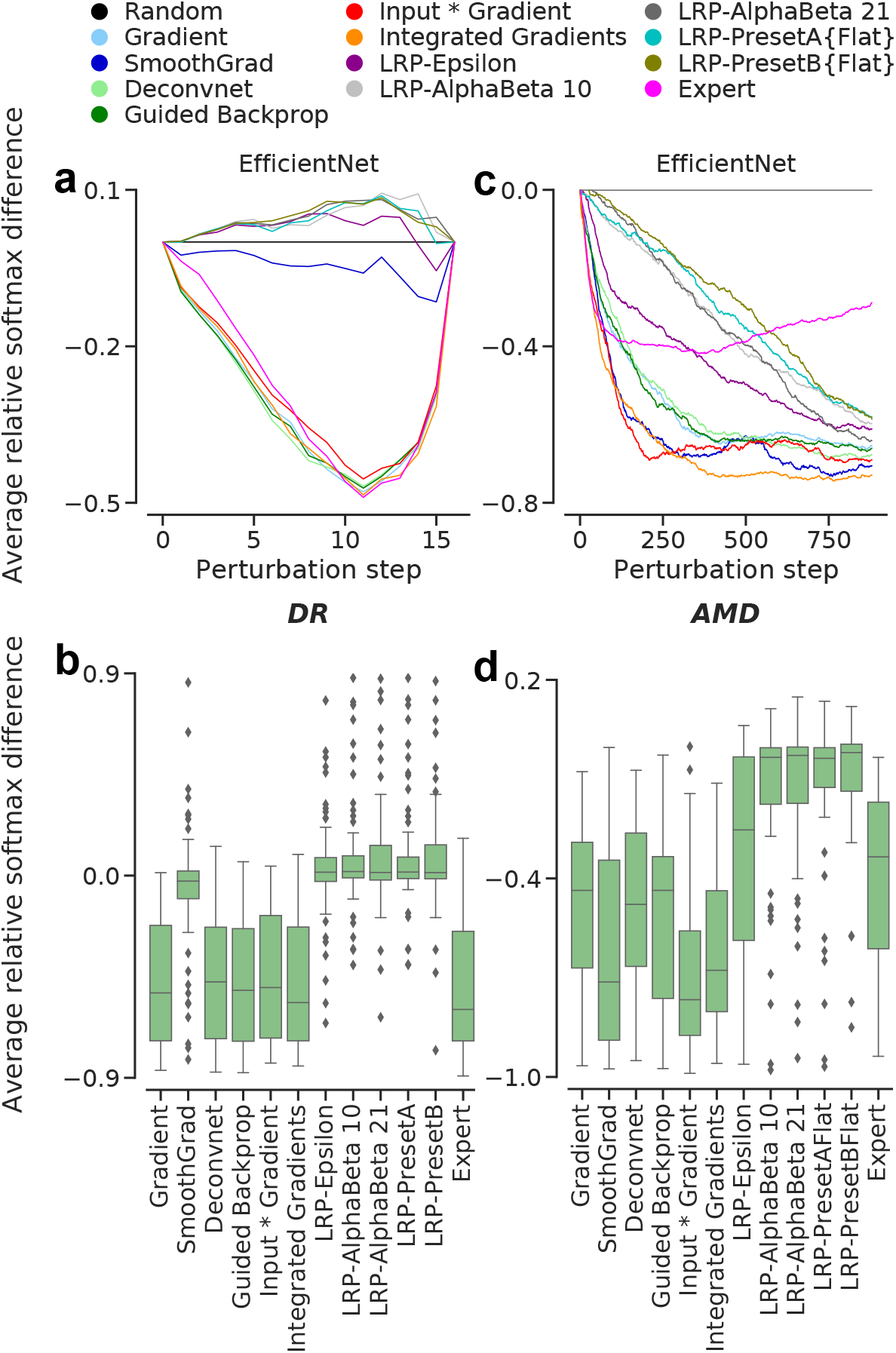
Perturbation analyses including the expert annotations as saliency maps. Curves were obtained by measuring the average differences from the *random* baseline. Thus, the baseline is shown as a flat line and all other methods converge to it, as the total perturbation grows and evidence is lost. Box plots show the distributions of the relative differences at steps 10 and 200, for the overall DR and nAMD scenarios, respectively. **(a-b)** Results for the DR detection task w.r.t. expert annotations *excluding the optic disc*. **(c-d)** Results for the nAMD activity detection task with complete expert annotations.

## Discussion

DR and nAMD are two progressive eye diseases and major causes of blindness in the developed world [18, 3, 95]. Timely intervention is the key to avoiding them or preventing vision loss in both cases. Thus, clinicians need cost-effective, accurate and trustworthy solutions to support the early diagnosis at scale [95, 35, 48, 20, 89]. Here, we developed accurate and well-calibrated ensembles of DNNs to detect DR and nAMD from retinal fundus images and slices of 3D OCT volume scans, respectively, and evaluated a comprehensive set of saliency maps for explaining the ensemble-based diagnostic decisions using a variety of published methods.

Interestingly, even the ensemble-based saliency maps were not readily interpretable by humans due to their sparsity. To improve the visualization of salient regions, we introduced a new post-processing method. Then, we systematically validated saliency maps against clinicians through two main analysis routes, including (1) a direct comparison of saliency maps with expert annotations of disease-specific pathologies and (2) perturbation analyses using also expert annotations as saliency maps. We found that the choice of DNN architecture and explanation method significantly influenced the quality of saliency maps. Moreover, DNNs used features both inside and outside the *regions-of-interest* (ROIs) annotated by clinicians. In particular, DNNs found additional instances of DR lesions that had not been explicitly annotated by clinicians. This could be because the heavily diseased images in the IDRiD dataset had not been completely annotated. In the nAMD case, extra cues were found in the fovea, which was never annotated by ophthalmologists in our study, as they only focused on signs of AMD activity they would typically use for diagnosis.

Saliency map generation to explain a classifier’s decision is superficially related to another popular task called semantic segmentation. However, segmentation is a *causal* task, while classification is *anti-causal* [17]. Also, DNNs are opportunistic classifiers in the sense that they exploit statistical regularities and image features to reach their objectives [31, 32]. Therefore, saliency maps for explaining the decisions of DNNs trained to achieve classification may differ from the segmentation maps typically used to train DNNs for segmentation in the first place. However, we gained insights into the diagnostic decisions of DNNs through the comparisons of saliency maps with expert annotations presented as segmentation maps. For instance, our DR detection networks mostly used a subset of small but sufficiently informative lesions, such as microaneurysms and hard exudates as well as small instances of hemorrhages. They also exploited soft exudates and large hemorrhages, albeit less frequently and only partially. Overall, they used efficient decision rules [32] mostly based on the characteristics of Mild and Moderate DR, as the task was to detect only the presence of DR. The opportunistic nature of DNNs also showed in *nAMD* activity detection. For instance, they detected large retinal fluid simply by its boundaries. Also, they exploited the fovea along with retinal fluid. Given that retinal fluid caused changes in the foveal contour during nAMD [57, 79], DNNs probably associated these changes with disease activity. Even though such associationist characteristics would not lead to causal explanations in principle [69], saliency maps showed that the DNN decisions were medically plausible. In this respect, DNNs, provided that they are also coupled with well-calibrated uncertainty estimation [10], can be deployed to facilitate the cooperation of clinicians and algorithms in the form of assisted reading [77]. Nevertheless, saliency maps are not recommended for lesion localization, especially not when one is interested in identifying all lesions, as also suggested by [12, 8].

In addition, our analyses indicated key practical limitations of the saliency methods in question. First, DR lesions such as microaneurysms and hard exudates as well as small bodies of retinal fluid in the case of nAMD indicate early-onset cases. As DNNs exploit retinal images opportunistically and the resulting saliency maps may include sparse regions even after our post-processing, the pitfall is that such minuscule but critical pathologies can be overlooked while screening for timely intervention. To alleviate this, alternative saliency methods designed for coarse maps can be used. Grad-CAM [78] and its combinations with Guided Backprop, or saliency bounding boxes [51] are good candidates to that end. Coarse maps can be also obtained from BagNets with *built-in* interpretability [15, 39]. Another important factor, which is somewhat neglected in our study, is the inter-grader variability in human readings of medical images. The inter-grader variability is high [24, 45], especially in segmentation tasks due to technical challenges and anatomical variability across patients [56]. Clinician performance is also subject to internal biases and experience levels. Thus, a more refined assessment of saliency maps could be achieved through multiple readers, also by estimating the ground truth segmentation from their annotations [98].

Our analyses on the DR detection task also excluded the optic disc annotations, while the optic disc can, of course, exhibit signs of DR, e.g., neovascularization. However, the relative size of neovascularizations were fairly small compared to the optic disc size. As the inclusion of the optic disc annotations would unnecessarily increase the Dice loss and complicate our analyses, we focused on the DR lesions outside the optic disc. Complementary figures including the optic disc can be found in Fig. 12 in Appendix A.3.

The decision mechanisms of DNNs and clinicians have also been recently compared via a perturbation-based reader study in the context of breast cancer screening [53]. The study included two groups of patients with either microcalcifications or soft tissue lesions, and indicated the bias of DNNs towards high-frequency features in both groups. While sharp and local peaks in mammogram images were salient features of microcalcifications, DNNs recognized soft tissue lesions typically from their boundaries without focusing on interiors. This is in line with our finding that the networks for DR and active nAMD detection used rather microaneurysms and the boundaries of large retinal fluid in the eye to make decisions. Also in line with our results, cancer screening networks found additional information outside the ROIs determined by radiologists [53].

In another recent study [81], an instance of InceptionV3 [86] was trained to predict the presence of choroidal neovascularization (CNV), diabetic macular edema (DME) or drusen from OCT images. Then, three experts graded saliency maps for its decisions on a scale between 0 and 5 according to their clinical relevance. In total, 13 saliency methods were used (9 of which are also used by our study). According to the subjective expert rating, Deep Taylor decomposition [61] and Guided Backprop [83] produced the most relevant saliency maps. Deep Taylor decomposition provided slightly better visualizations than Guided Backprop “*due to clinically coherent explanations, better coverage of pathology, and lack of high-frequency noise*” [81, p.7]. Thus, their study provides further evidence that Guided Backprop is a useful technique for obtaining clinically relevant saliency maps, especially considering that they did not use any special post-processing of the saliency maps for Guided Backprop (see Methods), which could have improved its saliency maps. Deep Taylor decomposition, however, performed less well in our study, hinting at a disagreement between their rating-based evaluation and our segmentation-based evaluation.

In contrast to this evidence by us and others [5, 81] in favor of Guided Backprob in a clinical setting, Guided Backprob has been shown to be insensitive to the object classes in ImageNet [75, 66]. This likely happens because the algorithm exploits local connections in convolutional layers, which extract a series of hierarchical feature representations from a given image, and the final dense layers, where class label assignments are made, have less impact on saliency maps [66]. Nevertheless, as Guided Backprop was consistently among the best methods for generating saliency maps to explain the decisions of DNNs trained to detect retinal diseases in our and other studies, we believe that it should be further studied to understand its distinct behaviors when explaining DNN decisions on natural or medical images. Moreover, its restriction by design to ReLU networks (see Methods) should be studied further to improve its applicability to new architectures beyond ReLU-based designs, such as EfficientNets [87, 88].

## Conclusion

We studied the clinical relevance of saliency maps extracted from DNNs trained to detect DR and nAMD from retinal images. We used different network architectures, well-calibrated ensembles of DNNs and a variety of explanation methods to obtain a comprehensive set ofsaliencymaps for explaining the ensemble-based diagnostic decisions. Then, we validated the saliency maps against ophthalmologist’s expert annotations. Overall, Guided Backprop emerged as the method of choice for generating saliency maps to explain the diagnostic decisions of DNNs on retinal images. In addition, a combination of multiple methods may reveal complementary characteristics in order to obtain well-rounded explanations.

## Data Availability

Retinal fundus images used in this study were obtained from publicly available repositories indicated in the manuscript.
The optical coherence tomography scans were obtained from the University Eye Clinic and their use was permitted by the Institutional Ethics Committee of the University of Tuebingen.

## Acknowledgements

This research was supported by the German Ministry of Science and Education (BMBF, 01GQ1601 and 01IS18039A) and the German Science Foundation (BE5601/4-1 and EXC 2064, project number 390727645). Additional funding was provided by Novartis AG through a research grant. The funders did not have any influence in the study planning and design. The Messidor 2 collection [21] was kindly provided by the Messidor program partners. More information can be found at http://www.adcis.net/en/third-party/messidor/.

## Author Contributions Statement

MSA and PB designed research; LBK devised the method for saliency map processing; MSA and LBK performed research, the AMD and DR experiments, respectively; GA gathered the OCT volumes and graded B-scans; WI also graded B-scans; LK annotated B-scans and provided medical advice together with GA, WI and FZ; MSA and PB supervised research; MSA, PB and LBK wrote the paper with input from all authors.

## Appendix

### A.1 Properties of *ν*(*τ*)

#### Lemma 1.

*ν*(*τ*) *is a monotonically decreasing and implicit function, where*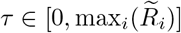.

*Proof*. Let *τ*_1_ *≤ τ*_2_, then

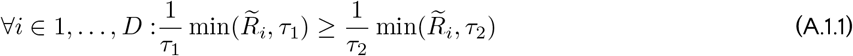

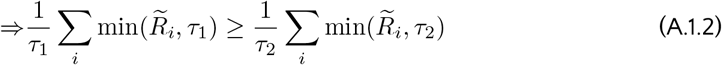

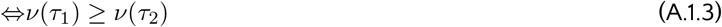

□

Given *τ*_1_ *≤ τ*_2_, A.1.1 holds true in the following cases:

- 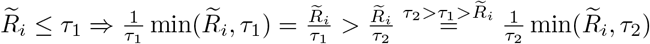
- 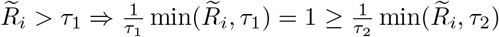

Then, *A*.1.1 ⇒ *A*.1.2 ⇔ *A*.1.3

### A.2 Optimum *f*_*ν*_ values for attribution methods and exemplary saliency maps

### A.3 Evaluation of saliency maps w.r.t. lesion types and their annotations

#### DR detection

#### AMD activity detection

### A.4 ANOVA and Post-hoc Tests

#### Direct comparison of saliency maps with expert annotations, DR detection

#### Direct comparison of saliency maps with expert annotations, AMD activity detection

#### Perturbation analysis, DR detection

#### Perturbation analysis, AMD activity detection

### A.5 Revisiting the disease detection performance and saliency maps evaluation with EfficientNets (B5)

## Footnotes

1 `np.geomspace(0.0005, 1.0, num=20, endpoint=True)` [67, 91])

